# Four-week forecasts of COVID-19 epidemic trajectories in South Africa, Chile, Peru and Brazil: a model evaluation

**DOI:** 10.1101/2021.09.06.21263151

**Authors:** Wim Delva, Emanuel M. Dominic, Roxanne Beauclair, Rachid Ouifki, Matthys Kroon, Heinrich Volmink, Elton Dorkin

## Abstract

**Introduction:** From the beginning of the COVID-19 pandemic, epidemiological models have been used in a number of ways to aid governments and organizations in efficient planning of resources and decision making. These models have elucidated important epidemiological transmission parameters, in addition to making short-term projections.

**Methods:** We constructed a compartmental mathematical model for the transmission, detection and prevention of SARS-CoV-2 infections for regions where Anglo American has mining operations. We fitted the model to publicly available data and used it to make short-term projections. Finally, we evaluated how the model performed by comparing short-term projections to actual confirmed cases, retrospectively.

**Findings:** The average forecast errors for four-week-ahead projections ranged between 1% and 8% in all the countries and regions considered in this study. All but one region had more than 75% of the true values falling within the range of four-week-ahead projections. The quality of the projections improved with time as expected due to increased historical data.

**Conclusion:** Our model produced four-week forecasts with a sufficiently high level of accuracy to guide operational and strategic planning for business continuity and COVID-19 responses in Anglo American mining sites.

## Introduction

Since December 2019, the COVID-19 pandemic has done untold damage to the health of individuals, healthcare systems, and economies across the world. As of 23 August 2021, the WHO estimated over 211 million confirmed cases and 4.4 million deaths^1^. These numbers continue to compound daily as more infectious variants of the SARS-CoV-2 virus spread rapidly through populations. Thus, countries across the world have adopted different measures to mitigate the spread of infection, and prevent an overload of the health care systems. A variety of non-pharmaceutical interventions (NPIs) that have been proposed and used encompass travel bans, domestic movement restrictions, social distancing, contact tracing of infected cases, self-isolation of symptomatic people, mandatory use of face-masks in public, and shielding of high-risk populations. Vaccines have been rolled out rapidly over the past 8 months, but vaccine coverage remains highly unequal across countries, with poorer countries in the global South lagging far behind^2^.

From the beginning of the pandemic, epidemiological models have been used in a number of ways to aid governments and organizations in efficient planning of resources and decision making^3,4^. They have been instrumental in the comparison of the relative impacts of different interventions (e.g. social distancing, restrictions on air travel, school closures, use of face masks, etc.), on the trajectory of the epidemic^3,5–10^. More recently, models have explored a wide range of biological and behavioural aspects of COVID-19 vaccination programmes, including vaccine efficacy, emerging variants, social determinants of vaccine access and uptake, waning immunity, and changes in behaviour due to vaccination impact^11–15^. Models have elucidated important epidemiological transmission parameters, and have produced short-term projections of cases, hospitalizations, and deaths that can be expected in different populations^16–19^.

Despite the usefulness of epidemiological models, it can be difficult to make accurate predictions^20,21^. Forecasting relies on the use of accurate data about the population for which it is making projections in order to calibrate the model^16^. Some problems modellers face with regard to data quality range from under-diagnosis of COVID-19 cases, and delays in reporting of case counts, hospitalizations, and deaths^21^. Moreover, it can be difficult to gain access to key data such as average duration of infection and length of stay in hospital for the populations being studied. There is also a trade-off to be made between having too simplistic and too complex models. If you have models that are overly simple, the predictions may be invalid because they have not captured the full dynamics of biology (e.g. virus evolution) and behaviour of individuals in a population (e.g. societal norms changing through time in regards to mask wearing). When models are too complex, they may become overly sensitive to small changes in parameters that are context-specific and difficult to estimate accurately^16^.

COVID-19 models, in particular, have problems with non-identifiability. Since there is still a paucity of publicly available data to calibrate models to, modellers tend to fit their data to confirmed cases and deaths only. The problem with this, is that there are many combinations of parameters that can produce a good fit to the data, yet have drastically different projected results^22,23^. Thus, it is often difficult to know if predictions are accurate, even if the model fits the available data well.

We constructed a mathematical model to assist Anglo American – a leading multinational mining company with operations in over 50 countries in the world and over 90 000 permanent employees worldwide^24^ –with their efforts to minimize the spread of COVID-19 among their workforce. We developed separate spatially homogeneous models for countries where Anglo American has most of their mining operations - Brazil, Peru, Chile, and South Africa. Additionally, we also constructed separate models for three South African provinces: Limpopo, Northern Cape and North West. The models were calibrated to publicly available data on laboratory-confirmed SARS-CoV-2 infections and deaths in these regions, and then four-week projections were made. Our primary objective here was to evaluate how the model performed when comparing short-term projections to actual confirmed cases, retrospectively.

## Methods

### Model structure and flow

We developed a compartmental model for the transmission and detection of SARS-CoV-2 infections. Figure 1 shows the health states and possible transitions between them. The model assumes that all people except for the seed infections are initially susceptible to SARS-CoV-2 infection. Upon infection, they transition to become “Exposed (*E*_*u*_)” but not yet infectious and not yet symptomatic. After a few days, they transition to the pre-symptomatic state (*I*_1*u*_) during which they are still asymptomatic, but now infectious, albeit less than during the next state of infection. From the pre-symptomatic state they may transition to either *I*_2*u*_, *I*_*mu*_ or *I*_*su*_. In these three states, they are respectively asymptomatic, mildly symptomatic, or severely symptomatic and very ill. People in the former two states will recover without needing hospital care. Severely ill patients, however, will either die (*D*_*s*_) before they reach a hospital, or they will be hospitalised (*H*). Diagnosis of SARS-CoV-2 infection can happen while in any of the infection states, but is more likely for symptomatic and severely symptomatic patients - in the absence of a contact tracing programme. Three events may occur among hospitalised patients: they may recover and be discharged (*R*_*h*_), they may die (*D*_*h*_) or they may require an admission to a critical care unit (*C*). The rate of hospitalisation and admission to critical care is not capped. People may leave the critical care unit either because they die (*D*_*c*_) or they are moved into a Post Critical care hospital (*P*) ward, from which they will transition to the recovered state (*R*_*c*_). The model is formally represented by a system of differential equations, given in the Supplementary information section at the end of this article.

**Figure 1.**
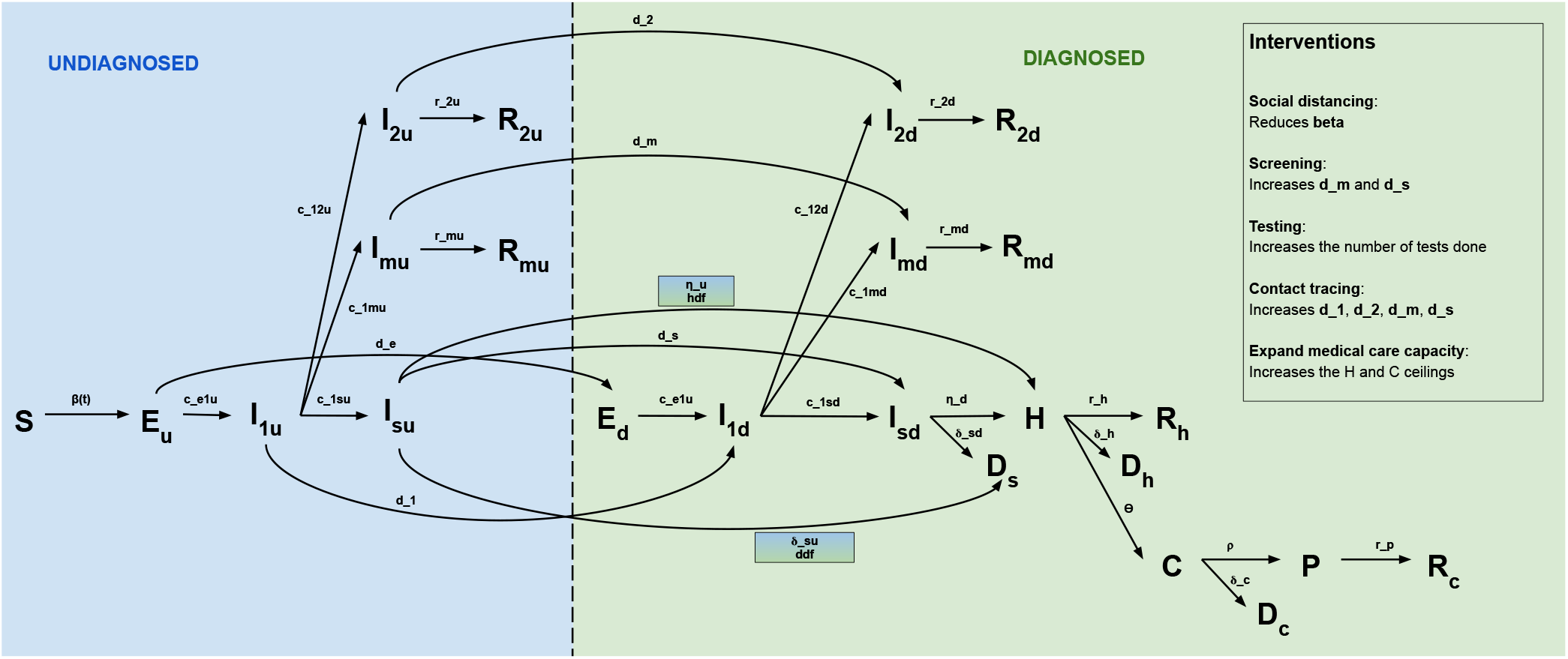
Schematic diagram of the disease transmission model. The capital letters represent different health states and the arrows represent transitions between the states. The letters above the arrows represents the rates of the transitions.

### Data

The model was calibrated to publicly available data (which we refer to as target features) on case counts and deaths. For the general South African population, and the South African provinces (Limpopo, Northern Cape and North West), we used data reported by the National Institute for Communicable Diseases (NICD) and the South African National Department of Health (DoH). The dataset was collated by the Data Science for Social Impact research group at the University of Pretoria^25^. For all the other regions (Brazil, Chile, Peru), we used data from Johns Hopkins University (JHU) Center for Systems Science and Engineering (CSSE) Github repository^26^. To ensure data consistency and quality, we compared these public data with data reported by Woldometer^27^.

### Model parameterization and calibration

Model parameters were informed by various epidemiological studies that estimated quantities (e.g. *r*_*mu*_; time from onset of symptoms to recovery for mildly symptomatic cases) based upon COVID-19 outbreaks in their respective populations. Most studies presented point estimates in the form of means and standard deviations or medians and inter-quartile ranges. Thus, we derived the bounds of the parameters’ prior distributions based upon descriptions of the spread of data around these point estimates. For some parameters (e.g. *r*_*md*_; time from diagnosis to recovery for mildly symptomatic cases), we did not have access to studies that directly quantified these particular parameters, and thus we made educated assumptions based on the priors for other parameters (e.g. *r*_*mu*_ and *d*_*m*_). Table 3 in the Supplementary information section at the end of this report shows the model parameters and table 4 gives a brief description of the parameters. The first seven parameters determine the effective contact rate between people susceptible to SARS-CoV-2 infection and people who are infected and infectious (in compartments *I*_1*\**_, *I*_2*\**_, *I*_*m\**_ or *I*_*s\**_), taking into account the seasonal variation in effective contact rates (*b*_*b*_). All the other parameters (excluding *h*_*ceil*_ and *c*_*ceil*_) determine the rate at which people transition between the health states in the model, as illustrated in Figure 1. Parameters *h*_*ceil*_ and *c*_*ceil*_ represent limits of the hospital and critical care capacity. For this modelling exercise, we kept the hospital and critical care limits uncapped by setting very large values for these two parameters. All these parameters drive the model’s features (i.e. cumulatively diagnosed SARS-CoV-2 infections and cumulative COVID-19 related deaths).

Informally, model calibration can be thought of as the process of searching for parameter values that produce model features that match the target features. We conducted a two-step calibration process. The first step involved manually adjusting the model parameters and then visually investigating the model fit. The second step involved using the model parameters from the first step and then applying a simple accept / reject Approximate Bayesian Computation scheme^28^. Sampling from the parameters’ prior distributions (a mix of unconditional uniform distributions and conditional distributions), the method filters through the explored parameter space and only retains parameter combinations that produce model features close to the target features. The distance between model features and target features was quantified by the relative Root-Mean-Squared-Error (RMSE), where the error terms were calculated by comparing the empirically observed time series of cumulatively diagnosed COVID-19 infections and cumulative deaths against the equivalent time series produced by the model. All time series of cumulative counts were rescaled by dividing all entries in the time series by the last (i.e. highest) value of the observed cumulative count. Due to high COVID-19 mortality underreporting that has been experienced in many regions^29^, we decided to give the our target features different weights during the calibration process. The cumulative cases time series was given a weight of 1 for all the regions. For the deaths time series, South Africa, Brazil, Peru, Chile and Limpopo were given a weight of 0.5 while Northern Cape and North West were given a weight of 0 (meaning that the model was not fitted to the deaths time series in these regions).

A total of one million parameter combinations were sampled from the prior distribution, and applied a tolerance threshold for the RMSE of 0.05. Thus, parameter combinations were retained if their associated model features stayed within 5% from the target features on average. We then narrowed down to get a more precisely calibrated model by setting the tolerance threshold for the RMSE at 0.001 and increasing this tolerance by 0.001 sequentially until at least forty parameter combinations were retained. The calibration scheme produced a posterior distribution for the parameters, consisting of multiple parameter combinations. This calibrated model was then used to generate projections of the number of COVID-19 deaths, active cases (stratified by severity), diagnosed and undiagnosed cases as well as the number of hospitalisations. In this paper, we only focus on projected COVID-19 cases. The model was re-calibrated once every month and the model projections updated.

### Model evaluation

We evaluated the performance of the model in making short term projections using frequently used summary metrics discussed below. To assess the difference between the model projections and the observed case counts, we computed the mean absolute percentage error (MAPE), a summary measure of the accuracy of forecasts. It expresses the error in the forecasts as a percentage. MAPE is defined as:

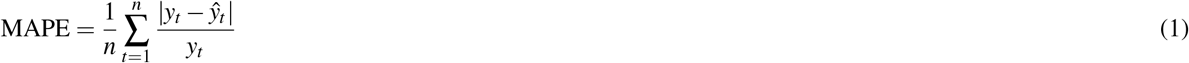

where *y*_*t*_ and *ŷ*_*t*_ denote the truth and the projected value at time *t* respectively, and *n* is the number of projections. The smaller the MAPE, the better.

To measure the of sharpness of the model, defined as the ability of the model to generate predictions within a narrow range of possible values, we used the median absolute deviation (MAD). MAD is a common statistic used to measure the spread out of a set of data. It is a property of the projections only and is defined as:

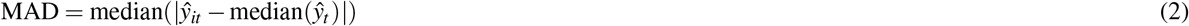

where *ŷ*_*it*_ denotes the projected value from each calibration at time *t*. A small MAD value means that the model is sharp while a large value means that the model is blurred.

We also computed the percentage of true values contained in the interquartile range and those contained between the range of all projections (between the minimum and maximum projections).

## Results

### COVID-19 projections

The calibrated models produced projections of cumulative cases of COVID-19. For all the regions, we conducted four-week ahead forecasts. Figure 2 shows the observed and predicted COVID-19 cases together with prediction intervals for different regions for each prediction period.

**Figure 2.**
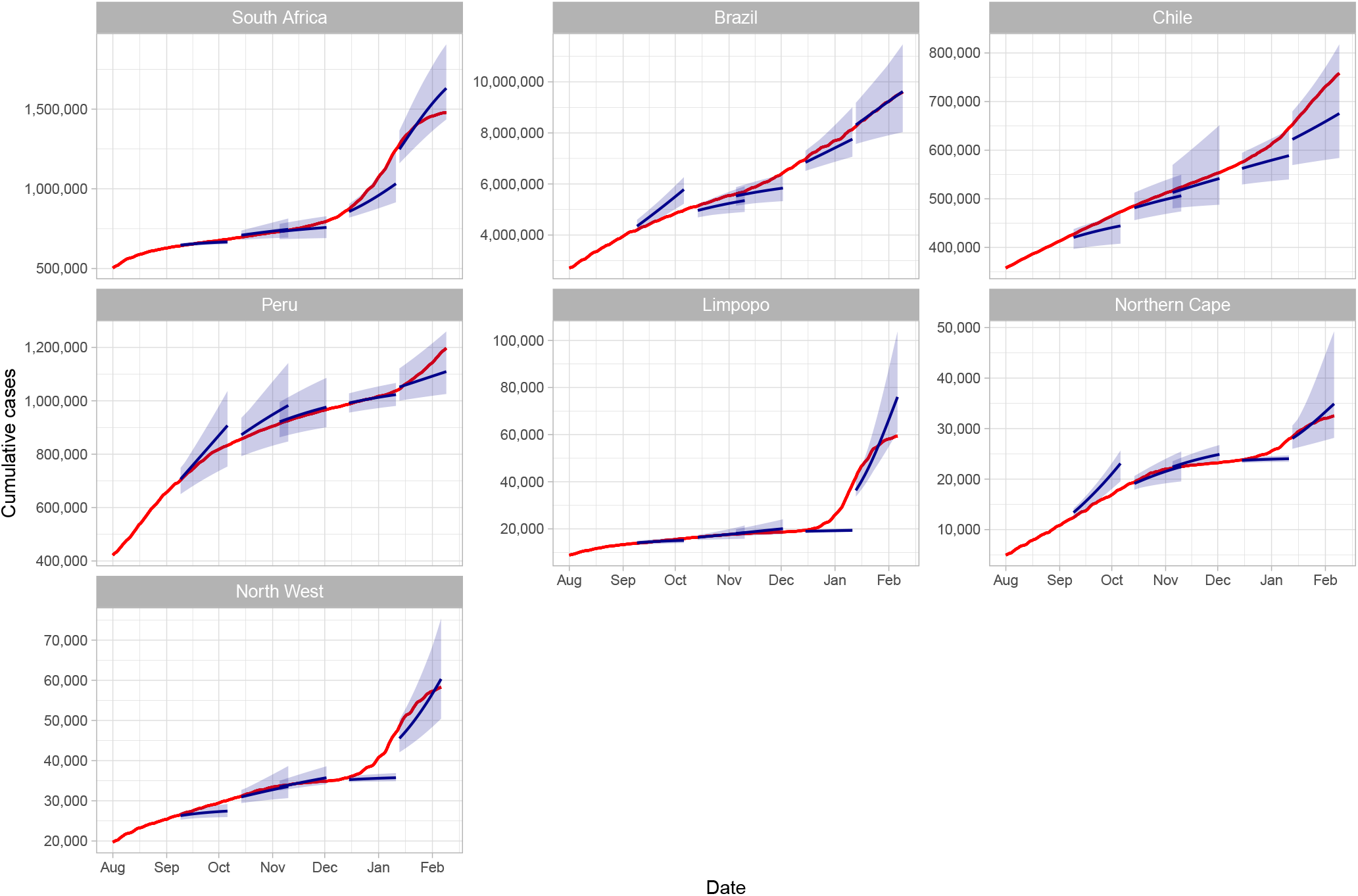
Cumulative COVID-19 confirmed cases together with forecasts and prediction intervals for different regions and origins (five time points) from September 2020 to January 2021. The red lines indicate the observed data and the blue lines indicate the projected mean cumulative cases. The shaded area indicates the minimum and maximum range.

The first forecast period covered September 9 - October 6, 2020. The average forecasts for South Africa and Limpopo were very close to the observed data and the range of projections produced were narrow. During the second and third forecast periods covering October 14 - November 10, 2020 and November 4 - December 2, 2020 respectively, the actual number of confirmed COVID-19 cases were contained in the prediction intervals in most of the regions. The prediction intervals were generally narrow as well. The model performed poorly in predicting a new wave of infection as seen in the case of South Africa during the fourth forecast period which covered December 14 - January 11, 2021. The model failed to accurately predict the increase in cases during the second wave of infections in December as seen in the plots for South Africa, Limpopo, Northern Cape and North West. The fifth forecast period covered January 13 - February 9, 2021. During this forecast period, the actual number of confirmed COVID-19 cases were contained in the prediction intervals in all of the regions. However, in some regions such as Chile and Limpopo, the mean estimates for the confirmed cases were far from the observed cases.

### Model evaluation

Figure 3 shows the overall model performance for difference regions. In all the regions, the forecast error increased as the forecast horizon increased. Nevertheless, the overall percentage forecast error for the four-week-ahead forecast was below 5% in all the regions besides Limpopo and Northern Cape. At one-week-ahead, most models had between 40% and 60% of the true values contained in the interquartile range. Peru had 100% of the true values contained in the interquartile range for one-week-ahead forecasts while Chile had the least number of true values falling within the interquartile range (35%). In most of the regions, the percentage of true values contained in the interquartile range decreased with time. All the regions had 60% or more of the true values falling within the range of model projections for the four-week-ahead projections. Brazil’s model was the most blurred model because it had the largest median absolute deviation values across the forecast horizon. This model exhibited rapid decrease in sharpness as the forecast horizon increased. Table 1 in the supplementary information section provides more information on overall performance of the model.

**Figure 3.**
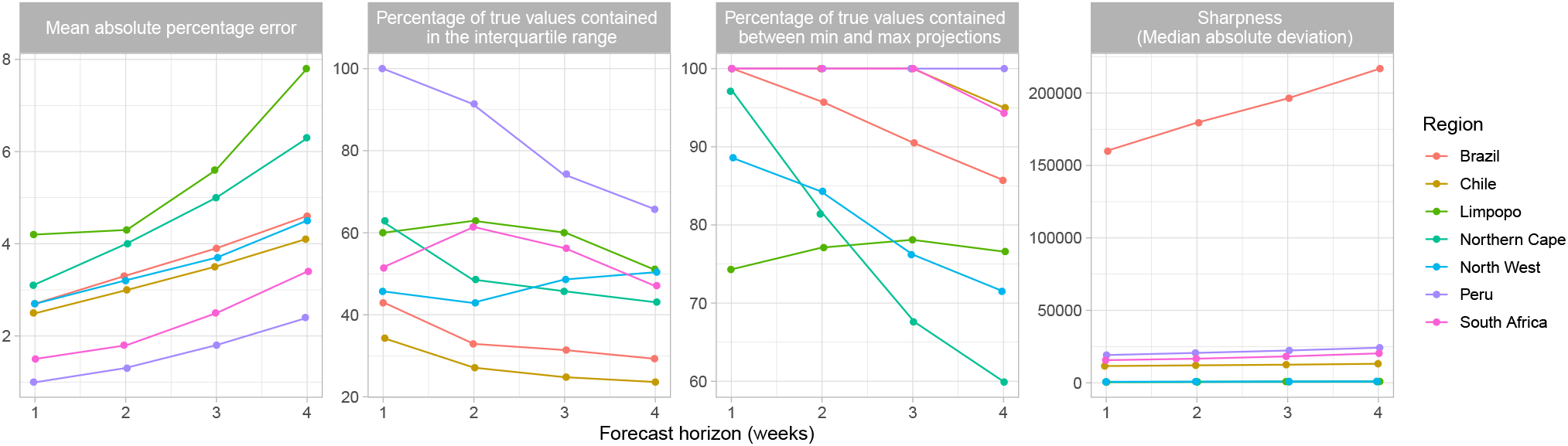
Overall model performance for the four week ahead forecasts across different regions.

Next, we analysed the model performance at different calibration time points for all the regions (Figure 4). The percentage forecast error for all the regions and all calibration time points was below 22%. In the first three rounds of forecasts, the forecast error was decreasing with time but then started increasing due to the second wave of infections. The percentage of true values contained in the interquartile range do not seem to have improved in subsequent forecasts, although there was variation in different regions. Table 2 in the supplementary information section shows the accuracy metrics for each region and calibration origin.

**Figure 4.**
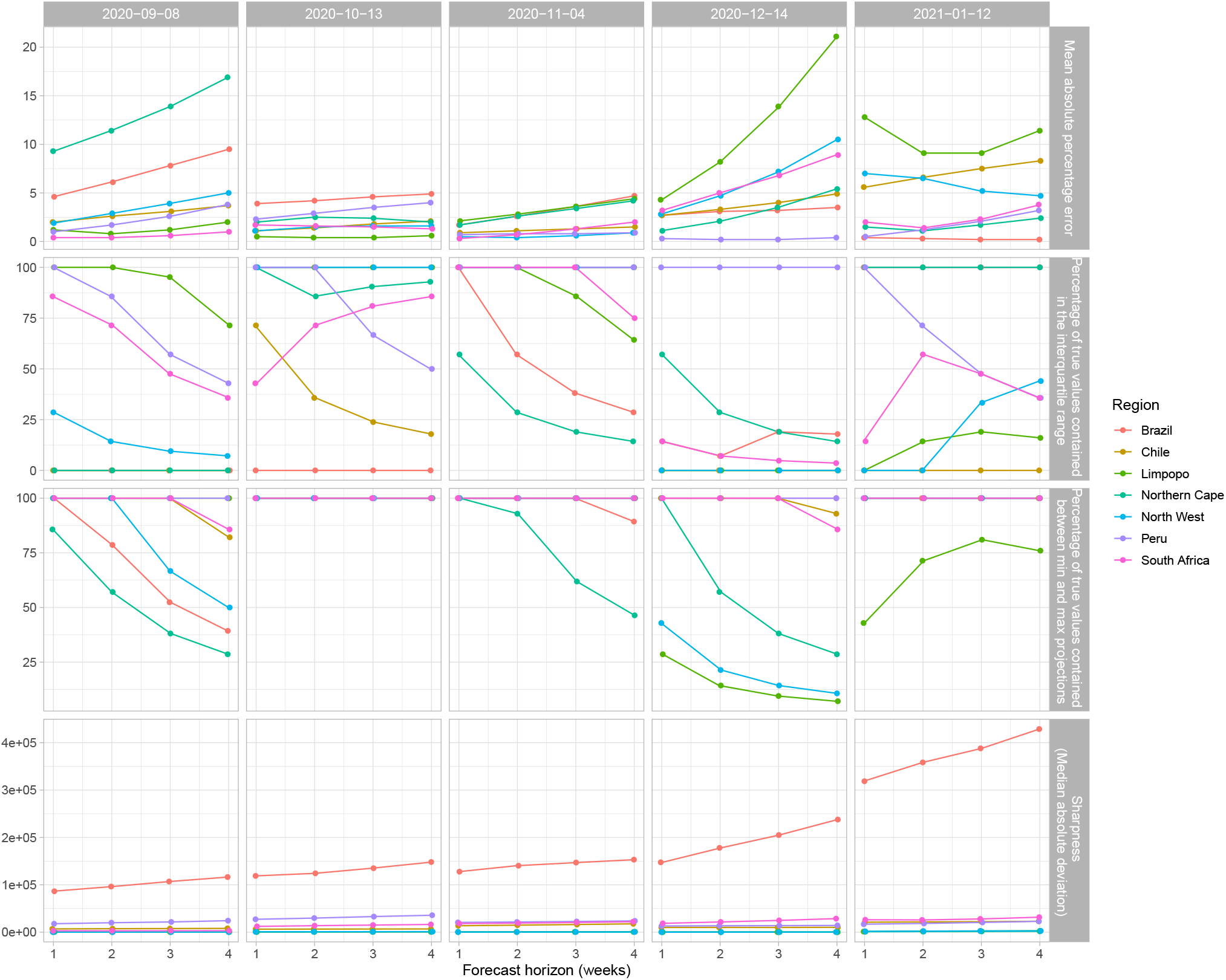
Model performance at each calibration time point.

## Discussion

The COVID-19 pandemic has continued to spread rapidly throughout the world. Several models have been developed to study the transmission dynamics and make short-term projections into the future about expected cases, hospitalisations and deaths^16–19^. We developed a compartmental model and calibrated it to publicly available data. We assumed that these data are accurate and reliable. The main objective here was to evaluate how the model performed when comparing short-term projections to actual confirmed cases, retrospectively.

Our model performed very well in producing short term projections with reported case counts falling within the range of projections. We obtained an overall forecast error of below 8% on average for four-week-ahead projections in all the countries and regions. As expected, the performance of the projections declined as the forecast horizon increased. This is because many processes that shape the epidemic continue to evolve as the disease evolves, most crucially processes at the molecular level (viral evolution, giving rise to viral variants with substantially different infectivity levels) as well as socio-political processs at the population level (government decisions to tighten or relax lockdown regulations). Neither of these sets of processes were explicitly captured by the model.

As the epidemic continues to evolve, more data will become available. The model’s parameter estimates will become more accurate, and the model-based inference will be less affected by parameter uncertainty. One such model-based epidemiological metric that may prove to be useful for continuous monitoring and evaluation of the COVID-19 response is the ratio of the moving averages of newly infected and newly recovered patients. An increase in this metric would indicate that the epidemic is getting worse, while a decrease would indicate the opposite.

Resurgence of the second wave of infections in different regions resulted in poor forecasts. Future improvements to the model involve redefining the effective contact rate 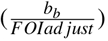. As lockdown regulations are loosened, we expect more contacts and hence high likelihood of increase in transmission.

Our model had an explicit observation process built-in to it. This was done for two reasons: firstly, it allowed us to calibrate the model to the available empirical data, since that data is based upon laboratory confirmed infections - which by definition exclude undiagnosed infections. Secondly, it will allow us to model the effect of interventions (e.g. amplified screening initiatives) that might change how many cases get diagnosed (i.e. what we observe in the real world). However, the challenge is that to date, there is very little literature on epidemiological parameters of those who are undiagnosed versus diagnosed. This influenced the uncertainties for relative infectiousness. For example, we assumed that being diagnosed likely makes a person relatively less infectious, because knowledge of infection would cause a case to modify their behaviour resulting in contact with fewer people through self-isolation. However, it is possible that diagnosed people with severe symptoms could be better at infecting others since those who remain undiagnosed likely have milder symptoms, which is suggestive of lower viral loads^30^. A related challenge is that we have almost no direct evidence to inform the various detection rates (parameters *d*_*e*_, *d*_1_, *d*_2_, *d*_*m*_ and *d*_*s*_) in the model. That in turn makes it virtually impossible to estimate what the impact of intensified contact tracing or screening might be. Relevant improvements in the South African SARS-CoV-2 surveillance system would therefore be immensely helpful in reducing parameter uncertainty and subsequently the uncertainty around model-based estimates and projections.

Our assumptions about the average time spent in each of the different compartments may also require future adjustments. Published estimates of duration spent in hospital or ICU, were not specific enough for our purposes. For instance, for *ρ* and *d*_*c*_, we used published estimates of length of stay in ICU. The estimate we based our priors on aggregated people together who may have died in ICU with those who were discharged back to a non-ICU hospital bed once they started to get better. Because of this, we made the assumption that those who don’t die have a longer time in ICU than those who do. This may affect how quickly all hospital and ICU beds fill up and how long they stay occupied, which in turn may affect the number total deaths resulting from the model.

The results presented here may also be influenced by the assumptions we made regarding disease severity, particularly in regards to the proportion of cases that remain completely asymptomatic throughout the duration of infection. It has been estimated that as many as 70% or more of infections^31,32^ could be asymptomatic. However, these studies do not follow-up cases past the full duration of the incubation period in order to see if symptoms develop later. Here, we assumed between 20-40% would be asymptomatic, based upon a systematic review of studies that estimated the proportion of asymptomatic cases when cases were followed-up for at least 14 days to determine their final symptoms status^33^. If asymptomatic cases are more common than we assumed, then we may expect fewer people to become cases in our model, since they are assumed to be less infectious than symptomatic cases.

In conclusion, even though limited data and considerable uncertainty around the transmission dynamics posed constraints to the accuracy and precision of our model forecasts, our model produced four-week forecasts with a sufficiently high level of accuracy to guide operational and strategic planning for business continuity and COVID-19 responses in Anglo American mining sites.

Several aspects of the model are being revised, in response to emerging knowledge that was not available at the start of the model development. In particular, waning immunity after naturally acquired SARS-CoV-2 infection, and the increasing coverage of COVID vaccines, yet with imperfect protection against infection, are important new features that are being added to the model. With respect to model calibration, comparison of smoothed time series of daily new cases and deaths rather than cumulative cases and deaths is being implemented.

## Data Availability

All data referred to in the manuscript are publicly available.
https://github.com/dsfsi/covid19za Marivate, V. & Combrink, H. M. Use of available data to inform the covid-19 outbreak in south africa: A case study. DataSci. J.19, 1-7 (2020).
https://github.com/CSSEGISandData/COVID-19 Dong, E., Du, H. & Gardner, L. An interactive web-based dashboard to track covid-19 in real time. The Lancet infectious diseases 20, 533-534 (2020).
https://www.worldometers.info/coronavirus/ Worldometer covid-19 coronavirus pandemic (2020).

## Supplementary Information

### Model performance

**Table 1.**
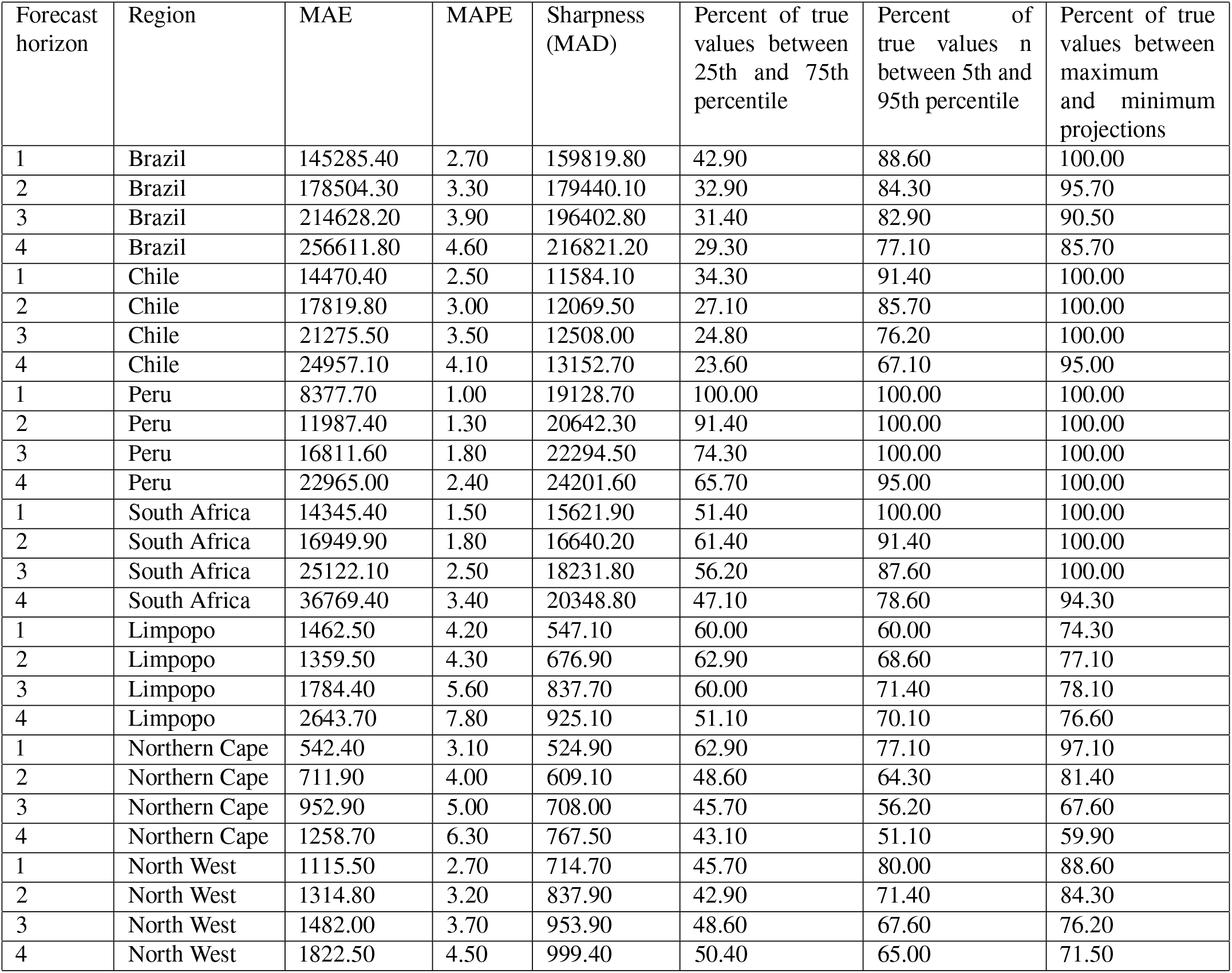
Overall model performance

**Table 2.** Model performance at different time points.

| Forecast horizon | Region | Calibration date | MAE | MAPE | Sharpness (MAD) | Percent of true values between 25th and 75th percentile | Percent of true values between 5th and 95th percentile | Percent of true values between maximum and minimum projections |
| --- | --- | --- | --- | --- | --- | --- | --- | --- |
| 1 | Brazil | 2020-09-08 | 198553.50 | 4.60 | 86391.50 | 0.00 | 42.90 | 100.00 |
| 2 | Brazil | 2020-09-08 | 271178.60 | 6.10 | 96282.90 | 0.00 | 21.40 | 78.60 |
| 3 | Brazil | 2020-09-08 | 354965.20 | 7.80 | 106962.60 | 0.00 | 14.30 | 52.40 |
| 4 | Brazil | 2020-09-08 | 446591.10 | 9.50 | 116356.60 | 0.00 | 10.70 | 39.30 |
| 1 | Brazil | 2020-10-13 | 200928.60 | 3.90 | 118789.90 | 0.00 | 100.00 | 100.00 |
| 2 | Brazil | 2020-10-13 | 223836.30 | 4.20 | 124242.00 | 0.00 | 100.00 | 100.00 |
| 3 | Brazil | 2020-10-13 | 249674.10 | 4.60 | 135053.40 | 0.00 | 100.00 | 100.00 |
| 4 | Brazil | 2020-10-13 | 270493.40 | 4.90 | 148113.70 | 0.00 | 100.00 | 100.00 |
| 1 | Brazil | 2020-11-04 | 98213.90 | 1.70 | 127708.70 | 100.00 | 100.00 | 100.00 |
| 2 | Brazil | 2020-11-04 | 150545.80 | 2.60 | 140312.60 | 57.10 | 100.00 | 100.00 |
| 3 | Brazil | 2020-11-04 | 211105.20 | 3.60 | 146982.20 | 38.10 | 100.00 | 100.00 |
| 4 | Brazil | 2020-11-04 | 284780.80 | 4.70 | 153138.40 | 28.60 | 75.00 | 89.30 |
| 1 | Brazil | 2020-12-14 | 196628.60 | 2.70 | 147524.50 | 14.30 | 100.00 | 100.00 |
| 2 | Brazil | 2020-12-14 | 223663.00 | 3.10 | 177999.00 | 7.10 | 100.00 | 100.00 |
| 3 | Brazil | 2020-12-14 | 237313.80 | 3.20 | 205022.40 | 19.00 | 100.00 | 100.00 |
| 4 | Brazil | 2020-12-14 | 263705.90 | 3.50 | 237683.80 | 17.90 | 100.00 | 100.00 |
| 1 | Brazil | 2021-01-12 | 32102.60 | 0.40 | 318684.20 | 100.00 | 100.00 | 100.00 |
| 2 | Brazil | 2021-01-12 | 23298.00 | 0.30 | 358363.90 | 100.00 | 100.00 | 100.00 |
| 3 | Brazil | 2021-01-12 | 20082.50 | 0.20 | 387993.30 | 100.00 | 100.00 | 100.00 |
| 4 | Brazil | 2021-01-12 | 17487.70 | 0.20 | 428813.60 | 100.00 | 100.00 | 100.00 |
| 1 | Chile | 2020-09-08 | 8877.90 | 2.00 | 6917.50 | 0.00 | 100.00 | 100.00 |
| 2 | Chile | 2020-09-08 | 11315.50 | 2.60 | 7354.50 | 0.00 | 100.00 | 100.00 |
| 3 | Chile | 2020-09-08 | 13900.60 | 3.10 | 7656.50 | 0.00 | 81.00 | 100.00 |
| 4 | Chile | 2020-09-08 | 16963.60 | 3.70 | 8012.50 | 0.00 | 60.70 | 82.10 |
| 1 | Chile | 2020-10-13 | 5428.10 | 1.10 | 6361.50 | 71.40 | 100.00 | 100.00 |
| 2 | Chile | 2020-10-13 | 7156.60 | 1.40 | 6494.90 | 35.70 | 100.00 | 100.00 |
| 3 | Chile | 2020-10-13 | 8847.60 | 1.80 | 6687.80 | 23.80 | 100.00 | 100.00 |
| 4 | Chile | 2020-10-13 | 10637.40 | 2.10 | 6817.20 | 17.90 | 100.00 | 100.00 |
| 1 | Chile | 2020-11-04 | 4905.50 | 0.90 | 13627.80 | 100.00 | 100.00 | 100.00 |
| 2 | Chile | 2020-11-04 | 5874.90 | 1.10 | 14816.00 | 100.00 | 100.00 | 100.00 |
| 3 | Chile | 2020-11-04 | 6865.90 | 1.30 | 16122.80 | 100.00 | 100.00 | 100.00 |
| 4 | Chile | 2020-11-04 | 7992.30 | 1.50 | 17662.20 | 100.00 | 100.00 | 100.00 |
| 1 | Chile | 2020-12-14 | 15748.00 | 2.70 | 9973.40 | 0.00 | 100.00 | 100.00 |
| 2 | Chile | 2020-12-14 | 19508.30 | 3.30 | 10043.20 | 0.00 | 100.00 | 100.00 |
| 3 | Chile | 2020-12-14 | 24046.40 | 4.00 | 9961.10 | 0.00 | 81.00 | 100.00 |
| 4 | Chile | 2020-12-14 | 30005.30 | 4.90 | 10211.80 | 0.00 | 60.70 | 92.90 |
| 1 | Chile | 2021-01-12 | 37392.40 | 5.60 | 21040.30 | 0.00 | 57.10 | 100.00 |
| 2 | Chile | 2021-01-12 | 45243.90 | 6.60 | 21639.00 | 0.00 | 28.60 | 100.00 |
| 3 | Chile | 2021-01-12 | 52717.20 | 7.50 | 22112.10 | 0.00 | 19.00 | 100.00 |
| 4 | Chile | 2021-01-12 | 59187.20 | 8.30 | 23059.90 | 0.00 | 14.30 | 100.00 |
| 1 | Peru | 2020-09-08 | 7062.70 | 1.00 | 17791.90 | 100.00 | 100.00 | 100.00 |
| 2 | peru | 2020-09-08 | 13105.20 | 1.70 | 19989.90 | 85.70 | 100.00 | 100.00 |
| 3 | Peru | 2020-09-08 | 20023.00 | 2.60 | 21574.60 | 57.10 | 100.00 | 100.00 |
| 4 | Peru | 2020-09-08 | 30146.60 | 3.80 | 24286.90 | 42.90 | 100.00 | 100.00 |
| 1 | Peru | 2020-10-13 | 20338.60 | 2.30 | 27241.90 | 100.00 | 100.00 | 100.00 |
| 2 | Peru | 2020-10-13 | 25354.20 | 2.90 | 29797.10 | 100.00 | 100.00 | 100.00 |
| 3 | Peru | 2020-10-13 | 30931.90 | 3.50 | 32880.20 | 66.70 | 100.00 | 100.00 |
| 4 | Peru | 2020-10-13 | 36204.00 | 4.00 | 35779.60 | 50.00 | 100.00 | 100.00 |
| 1 | Peru | 2020-11-04 | 6822.10 | 0.70 | 20618.80 | 100.00 | 100.00 | 100.00 |
| 2 | Peru | 2020-11-04 | 7315.60 | 0.80 | 21484.40 | 100.00 | 100.00 | 100.00 |
| 3 | Peru | 2020-11-04 | 7927.40 | 0.80 | 22567.90 | 100.00 | 100.00 | 100.00 |
| 4 | Peru | 2020-11-04 | 8293.60 | 0.90 | 23753.10 | 100.00 | 100.00 | 100.00 |
| 1 | Peru | 2020-12-14 | 2555.80 | 0.30 | 13161.80 | 100.00 | 100.00 | 100.00 |
| 2 | Peru | 2020-12-14 | 1490.50 | 0.20 | 13547.00 | 100.00 | 100.00 | 100.00 |
| 3 | Peru | 2020-12-14 | 2097.30 | 0.20 | 13849.80 | 100.00 | 100.00 | 100.00 |
| 4 | Peru | 2020-12-14 | 3923.50 | 0.40 | 14236.90 | 100.00 | 100.00 | 100.00 |
| 1 | Peru | 2021-01-12 | 5109.40 | 0.50 | 16829.10 | 100.00 | 100.00 | 100.00 |
| 2 | Peru | 2021-01-12 | 12671.30 | 1.20 | 18392.90 | 71.40 | 100.00 | 100.00 |
| 3 | Peru | 2021-01-12 | 23078.10 | 2.10 | 20599.80 | 47.60 | 100.00 | 100.00 |
| 4 | Peru | 2021-01-12 | 36257.10 | 3.20 | 22951.70 | 35.70 | 75.00 | 100.00 |
| 1 | South Africa | 2020-09-08 | 2476.50 | 0.40 | 2929.50 | 85.70 | 100.00 | 100.00 |
| 2 | South Africa | 2020-09-08 | 2425.80 | 0.40 | 2934.00 | 71.40 | 100.00 | 100.00 |
| 3 | South Africa | 2020-09-08 | 4146.80 | 0.60 | 3016.80 | 47.60 | 100.00 | 100.00 |
| 4 | South Africa | 2020-09-08 | 6589.50 | 1.00 | 3152.80 | 35.70 | 75.00 | 85.70 |
| 1 | South Africa | 2020-10-13 | 12150.00 | 1.70 | 12191.60 | 42.90 | 100.00 | 100.00 |
| 2 | South Africa | 2020-10-13 | 11204.80 | 1.60 | 13698.30 | 71.40 | 100.00 | 100.00 |
| 3 | South Africa | 2020-10-13 | 10329.10 | 1.50 | 14996.20 | 81.00 | 100.00 | 100.00 |
| 4 | South Africa | 2020-10-13 | 9416.40 | 1.30 | 16340.30 | 85.70 | 100.00 | 100.00 |
| 1 | South Africa | 2020-11-04 | 1985.60 | 0.30 | 18154.30 | 100.00 | 100.00 | 100.00 |
| 2 | South Africa | 2020-11-04 | 5104.50 | 0.70 | 19135.40 | 100.00 | 100.00 | 100.00 |
| 3 | South Africa | 2020-11-04 | 9531.30 | 1.30 | 20456.10 | 100.00 | 100.00 | 100.00 |
| 4 | South Africa | 2020-11-04 | 15210.40 | 2.00 | 21995.10 | 75.00 | 100.00 | 100.00 |
| 1 | South Africa | 2020-12-14 | 29367.60 | 3.20 | 18618.90 | 14.30 | 100.00 | 100.00 |
| 2 | South Africa | 2020-12-14 | 47809.80 | 5.00 | 21585.20 | 7.10 | 57.10 | 100.00 |
| 3 | South Africa | 2020-12-14 | 69065.50 | 6.80 | 24740.20 | 4.80 | 38.10 | 100.00 |
| 4 | South Africa | 2020-12-14 | 97055.40 | 8.90 | 28619.90 | 3.60 | 28.60 | 85.70 |
| 1 | South Africa | 2021-01-12 | 25747.10 | 2.00 | 26215.20 | 14.30 | 100.00 | 100.00 |
| 2 | South Africa | 2021-01-12 | 18204.60 | 1.40 | 25848.00 | 57.10 | 100.00 | 100.00 |
| 3 | South Africa | 2021-01-12 | 32537.90 | 2.30 | 27949.80 | 47.60 | 100.00 | 100.00 |
| 4 | South Africa | 2021-01-12 | 55575.40 | 3.80 | 31635.80 | 35.70 | 89.30 | 100.00 |
| 1 | Limpopo | 2020-09-08 | 171.30 | 1.20 | 326.90 | 100.00 | 100.00 | 100.00 |
| 2 | Limpopo | 2020-09-08 | 114.60 | 0.80 | 327.00 | 100.00 | 100.00 | 100.00 |
| 3 | Limpopo | 2020-09-08 | 182.20 | 1.20 | 329.90 | 95.20 | 100.00 | 100.00 |
| 4 | Limpopo | 2020-09-08 | 299.70 | 2.00 | 339.50 | 71.40 | 100.00 | 100.00 |
| 1 | Limpopo | 2020-10-13 | 82.90 | 0.50 | 434.30 | 100.00 | 100.00 | 100.00 |
| 2 | Limpopo | 2020-10-13 | 65.30 | 0.40 | 506.90 | 100.00 | 100.00 | 100.00 |
| 3 | Limpopo | 2020-10-13 | 65.50 | 0.40 | 586.20 | 100.00 | 100.00 | 100.00 |
| 4 | Limpopo | 2020-10-13 | 112.00 | 0.60 | 679.80 | 100.00 | 100.00 | 100.00 |
| 1 | Limpopo | 2020-11-04 | 381.00 | 2.10 | 567.20 | 100.00 | 100.00 | 100.00 |
| 2 | Limpopo | 2020-11-04 | 503.50 | 2.80 | 643.00 | 100.00 | 100.00 | 100.00 |
| 3 | Limpopo | 2020-11-04 | 653.00 | 3.60 | 731.10 | 85.70 | 100.00 | 100.00 |
| 4 | Limpopo | 2020-11-04 | 810.60 | 4.40 | 820.70 | 64.30 | 100.00 | 100.00 |
| 1 | Limpopo | 2020-12-14 | 848.80 | 4.30 | 175.00 | 0.00 | 0.00 | 28.60 |
| 2 | Limpopo | 2020-12-14 | 1765.90 | 8.20 | 187.80 | 0.00 | 0.00 | 14.30 |
| 3 | Limpopo | 2020-12-14 | 3387.20 | 13.90 | 194.00 | 0.00 | 0.00 | 9.50 |
| 4 | Limpopo | 2020-12-14 | 6228.50 | 21.10 | 202.10 | 0.00 | 0.00 | 7.10 |
| 1 | Limpopo | 2021-01-12 | 5828.70 | 12.80 | 1232.00 | 0.00 | 0.00 | 42.90 |
| 2 | Limpopo | 2021-01-12 | 4348.40 | 9.10 | 1720.00 | 14.30 | 42.90 | 71.40 |
| 3 | Limpopo | 2021-01-12 | 4634.10 | 9.10 | 2347.10 | 19.00 | 57.10 | 81.00 |
| 4 | Limpopo | 2021-01-12 | 6142.80 | 11.40 | 2782.60 | 16.00 | 48.00 | 76.00 |
| 1 | Northern Cape | 2020-09-08 | 1218.10 | 9.30 | 416.10 | 0.00 | 0.00 | 85.70 |
| 2 | Northern Cape | 2020-09-08 | 1602.60 | 11.40 | 447.00 | 0.00 | 0.00 | 57.10 |
| 3 | Northern Cape | 2020-09-08 | 2086.60 | 13.90 | 514.70 | 0.00 | 0.00 | 38.10 |
| 4 | Northern Cape | 2020-09-08 | 2683.00 | 16.90 | 606.30 | 0.00 | 0.00 | 28.60 |
| 1 | Northern Cape | 2020-10-13 | 401.20 | 2.00 | 590.80 | 100.00 | 100.00 | 100.00 |
| 2 | Northern Cape | 2020-10-13 | 516.20 | 2.50 | 657.10 | 85.70 | 100.00 | 100.00 |
| 3 | Northern Cape | 2020-10-13 | 502.50 | 2.40 | 724.20 | 90.50 | 100.00 | 100.00 |
| 4 | Northern Cape | 2020-10-13 | 417.20 | 2.00 | 800.10 | 92.90 | 100.00 | 100.00 |
| 1 | Northern Cape | 2020-11-04 | 383.60 | 1.70 | 327.30 | 57.10 | 100.00 | 100.00 |
| 2 | Northern Cape | 2020-11-04 | 582.50 | 2.60 | 374.70 | 28.60 | 78.60 | 92.90 |
| 3 | Northern Cape | 2020-11-04 | 779.60 | 3.40 | 408.30 | 19.00 | 52.40 | 61.90 |
| 4 | Northern Cape | 2020-11-04 | 961.50 | 4.20 | 443.20 | 14.30 | 39.30 | 46.40 |
| 1 | Northern Cape | 2020-12-14 | 255.80 | 1.10 | 237.50 | 57.10 | 85.70 | 100.00 |
| 2 | Northern Cape | 2020-12-14 | 517.90 | 2.10 | 243.00 | 28.60 | 42.90 | 57.10 |
| 3 | Northern Cape | 2020-12-14 | 872.50 | 3.50 | 251.40 | 19.00 | 28.60 | 38.10 |
| 4 | Northern Cape | 2020-12-14 | 1408.50 | 5.40 | 258.50 | 14.30 | 21.40 | 28.60 |
| 1 | Northern Cape | 2021-01-12 | 453.50 | 1.50 | 1052.60 | 100.00 | 100.00 | 100.00 |
| 2 | Northern Cape | 2021-01-12 | 340.40 | 1.10 | 1323.90 | 100.00 | 100.00 | 100.00 |
| 3 | Northern Cape | 2021-01-12 | 523.00 | 1.70 | 1641.40 | 100.00 | 100.00 | 100.00 |
| 4 | Northern Cape | 2021-01-12 | 771.20 | 2.40 | 1844.60 | 100.00 | 100.00 | 100.00 |
| 1 | North West | 2020-09-08 | 524.70 | 1.90 | 371.90 | 28.60 | 100.00 | 100.00 |
| 2 | North West | 2020-09-08 | 813.30 | 2.90 | 392.20 | 14.30 | 57.10 | 100.00 |
| 3 | North West | 2020-09-08 | 1118.10 | 3.90 | 404.00 | 9.50 | 38.10 | 66.70 |
| 4 | North West | 2020-09-08 | 1434.50 | 5.00 | 412.70 | 7.10 | 28.60 | 50.00 |
| 1 | North West | 2020-10-13 | 345.90 | 1.10 | 781.80 | 100.00 | 100.00 | 100.00 |
| 2 | North West | 2020-10-13 | 473.10 | 1.50 | 915.10 | 100.00 | 100.00 | 100.00 |
| 3 | North West | 2020-10-13 | 530.40 | 1.60 | 1039.30 | 100.00 | 100.00 | 100.00 |
| 4 | North West | 2020-10-13 | 524.70 | 1.60 | 1137.60 | 100.00 | 100.00 | 100.00 |
| 1 | North West | 2020-11-04 | 162.10 | 0.50 | 398.90 | 100.00 | 100.00 | 100.00 |
| 2 | North West | 2020-11-04 | 121.10 | 0.40 | 454.30 | 100.00 | 100.00 | 100.00 |
| 3 | North West | 2020-11-04 | 200.00 | 0.60 | 509.70 | 100.00 | 100.00 | 100.00 |
| 4 | North West | 2020-11-04 | 324.20 | 0.90 | 575.10 | 100.00 | 100.00 | 100.00 |
| 1 | North West | 2020-12-14 | 1012.40 | 2.80 | 210.70 | 0.00 | 0.00 | 42.90 |
| 2 | North West | 2020-12-14 | 1786.80 | 4.70 | 226.30 | 0.00 | 0.00 | 21.40 |
| 3 | North West | 2020-12-14 | 2835.70 | 7.20 | 240.10 | 0.00 | 0.00 | 14.30 |
| 4 | North West | 2020-12-14 | 4416.50 | 10.50 | 251.40 | 0.00 | 0.00 | 10.70 |
| 1 | North West | 2021-01-12 | 3532.20 | 7.00 | 1810.20 | 0.00 | 100.00 | 100.00 |
| 2 | North West | 2021-01-12 | 3379.80 | 6.50 | 2201.40 | 0.00 | 100.00 | 100.00 |
| 3 | North West | 2021-01-12 | 2726.00 | 5.20 | 2576.20 | 33.30 | 100.00 | 100.00 |
| 4 | North West | 2021-01-12 | 2483.60 | 4.70 | 2814.40 | 44.00 | 100.00 | 100.00 |

### Model parameters

**Table 3.**
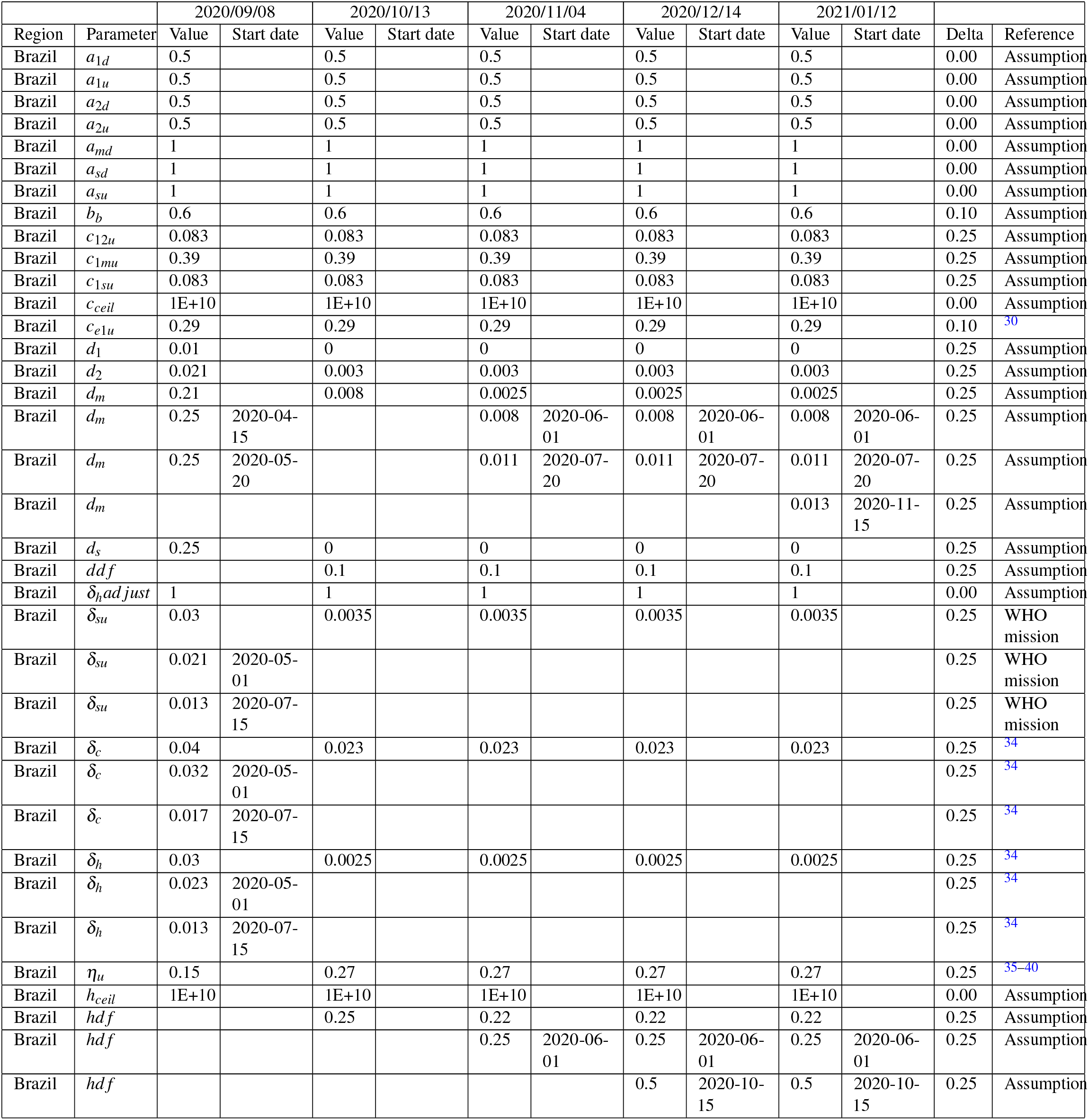
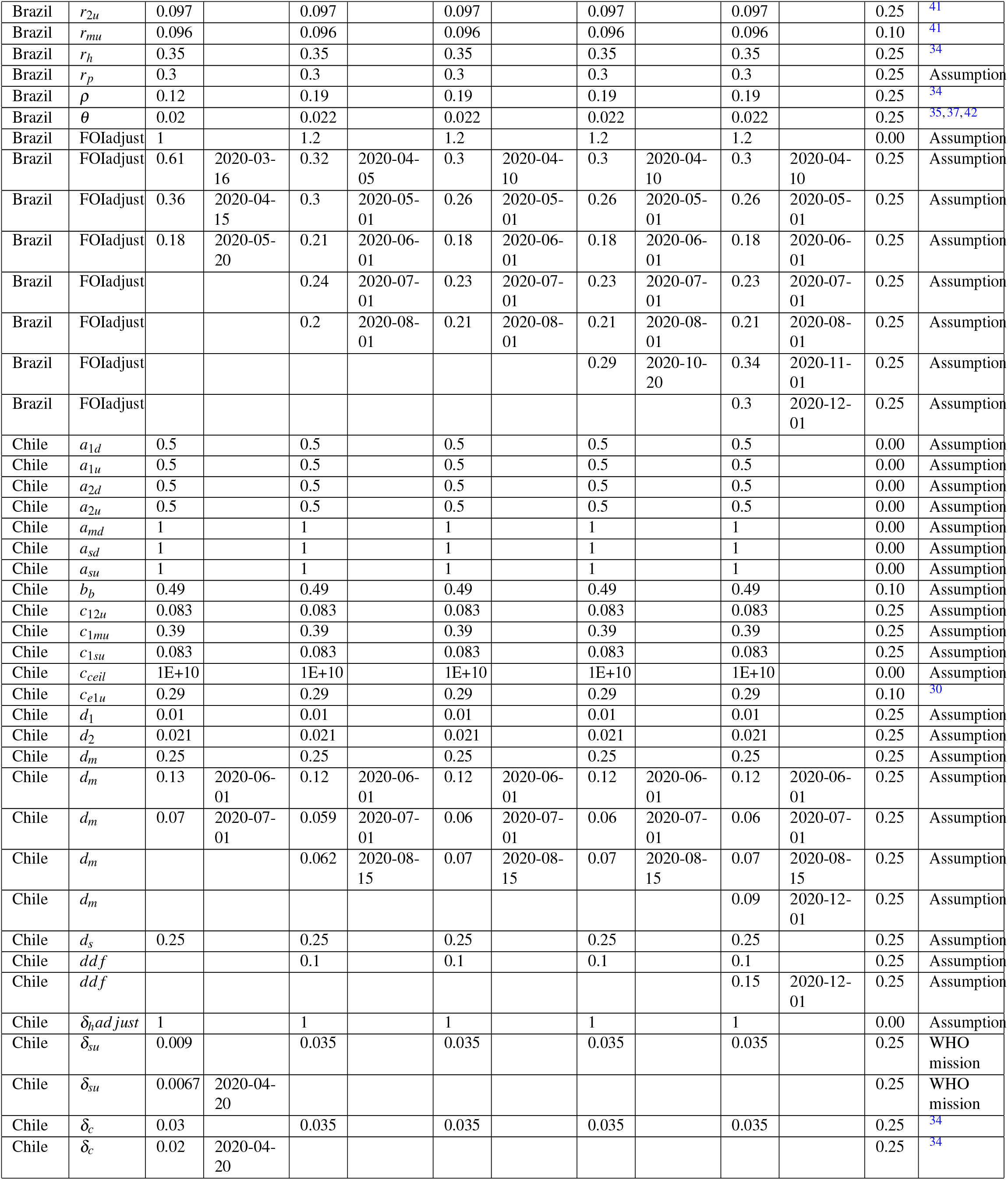
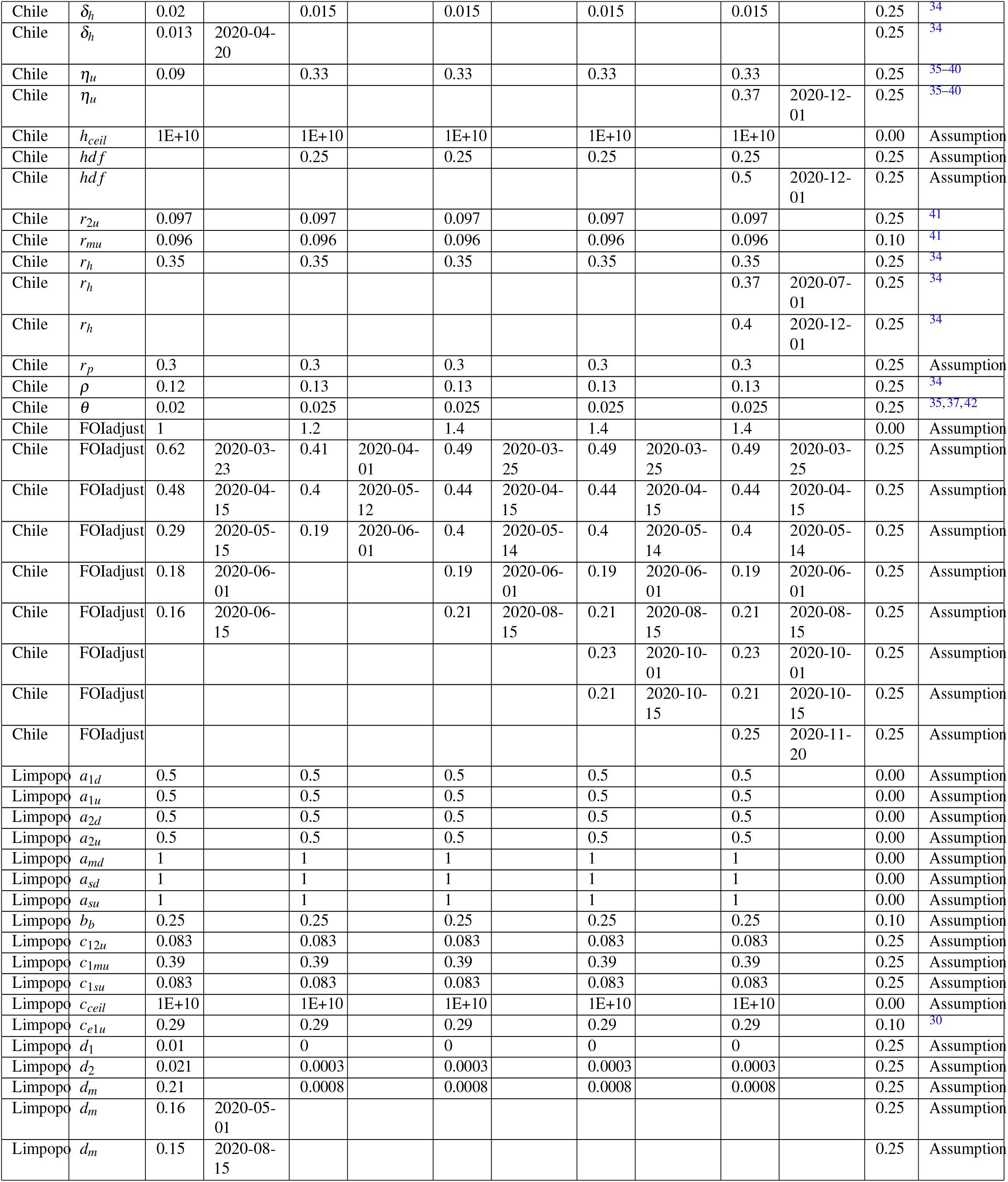
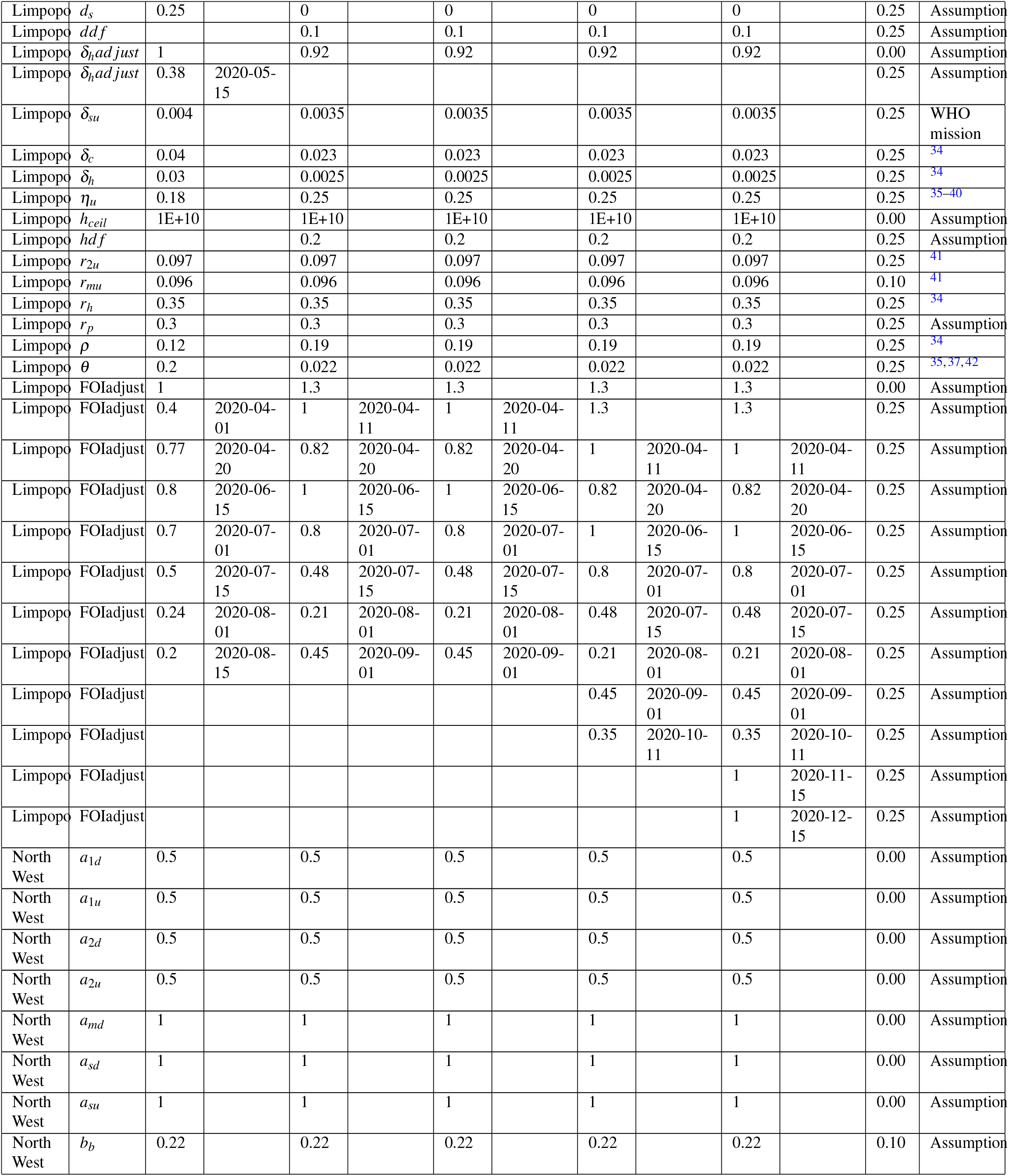
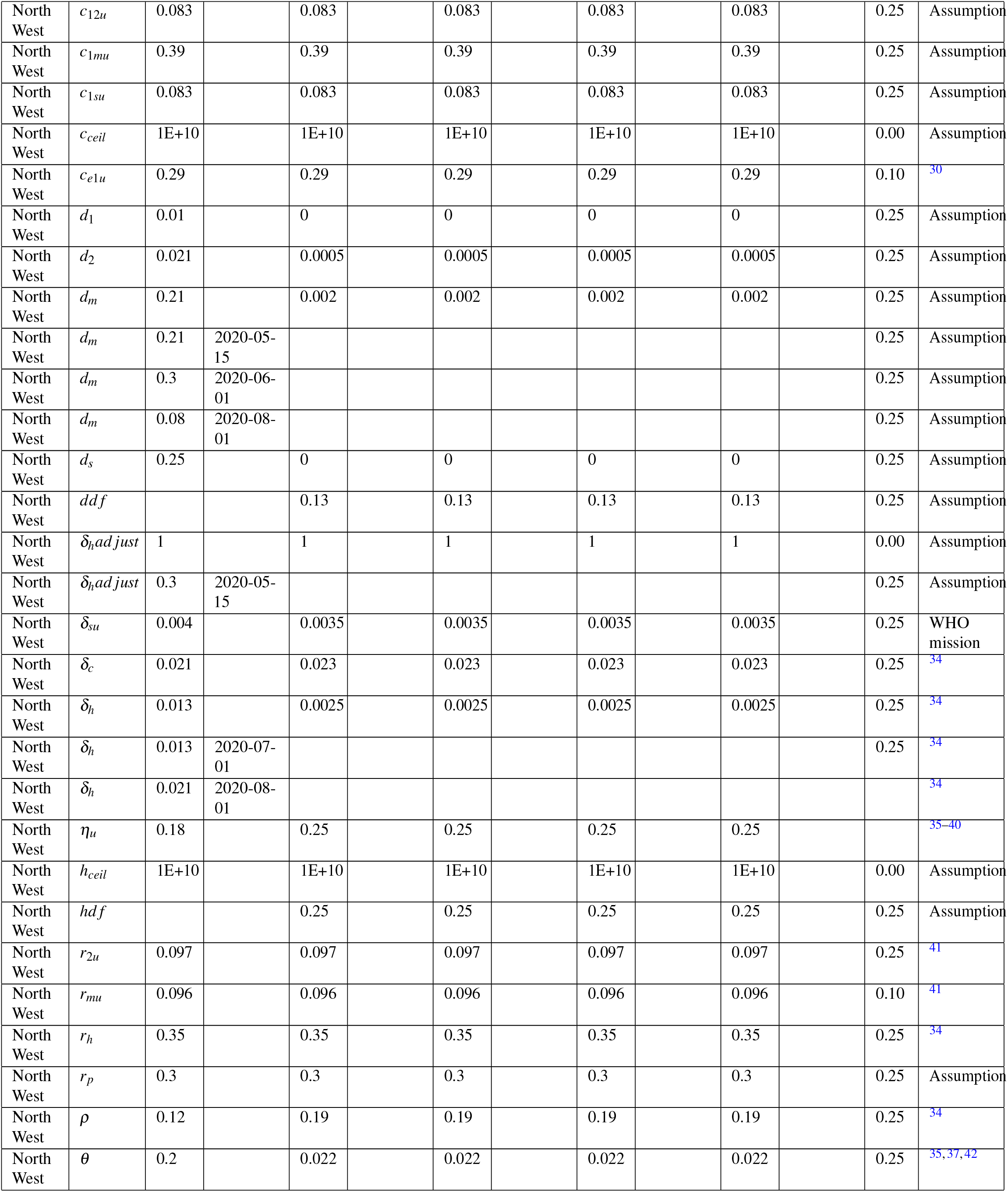
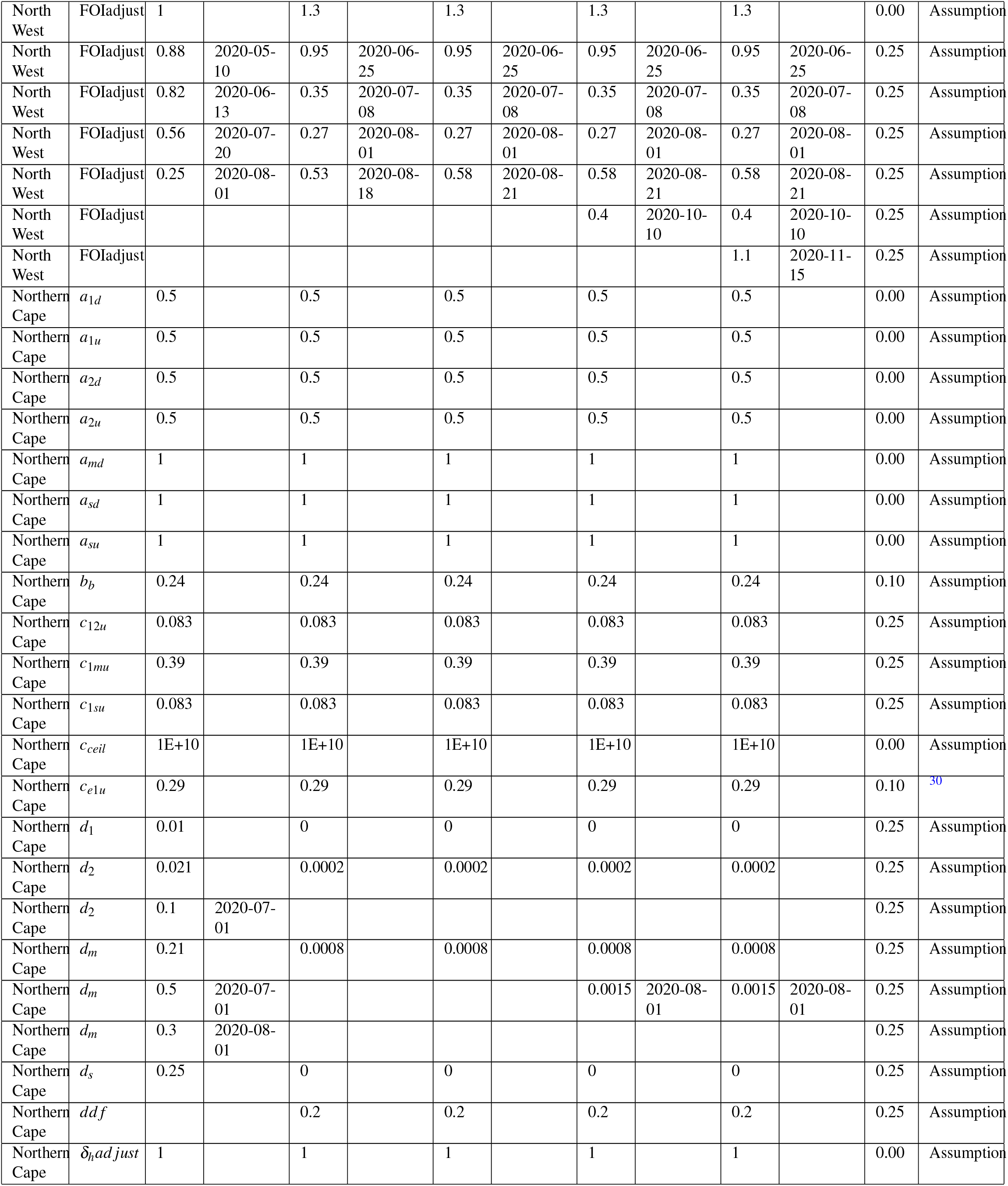
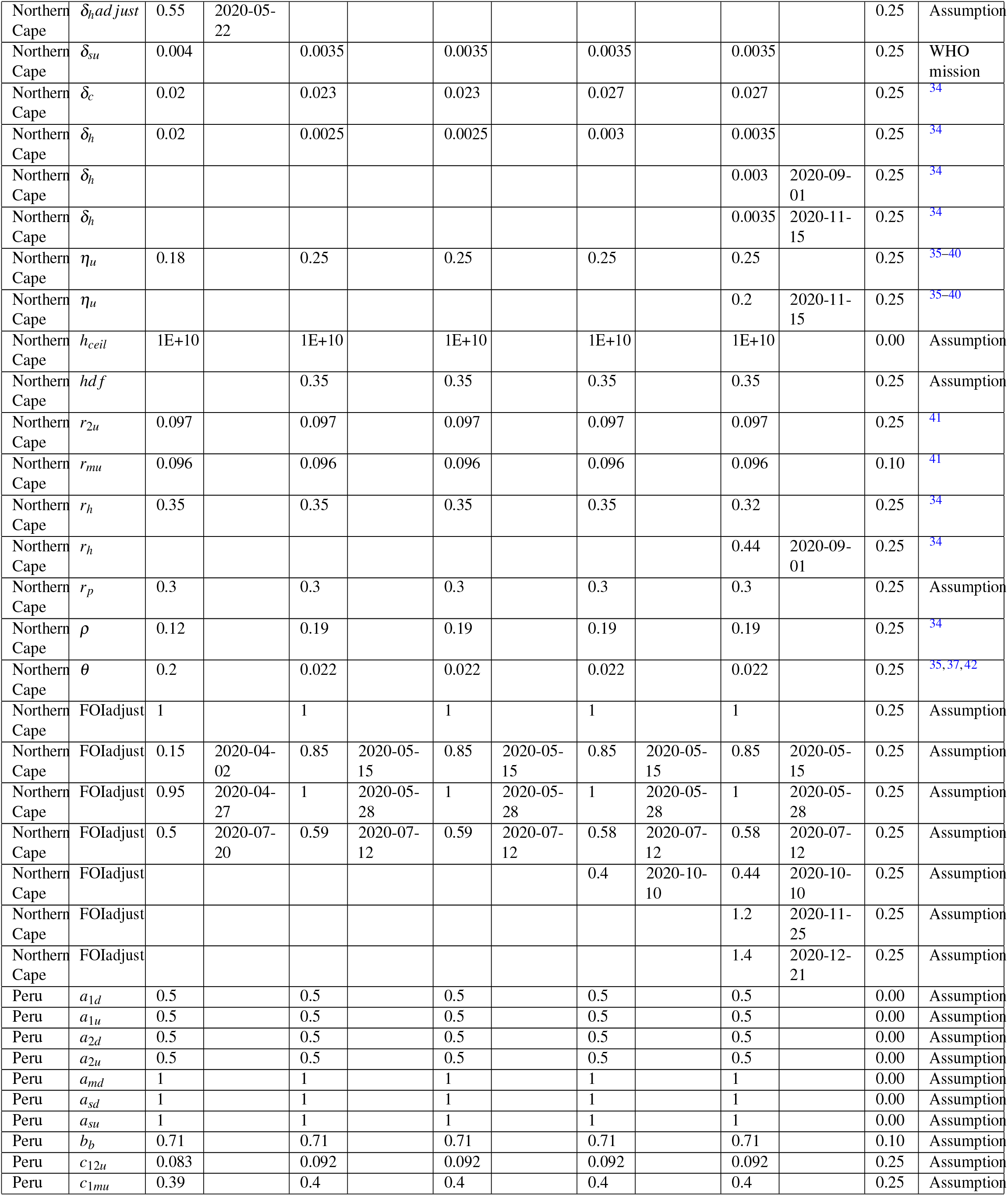
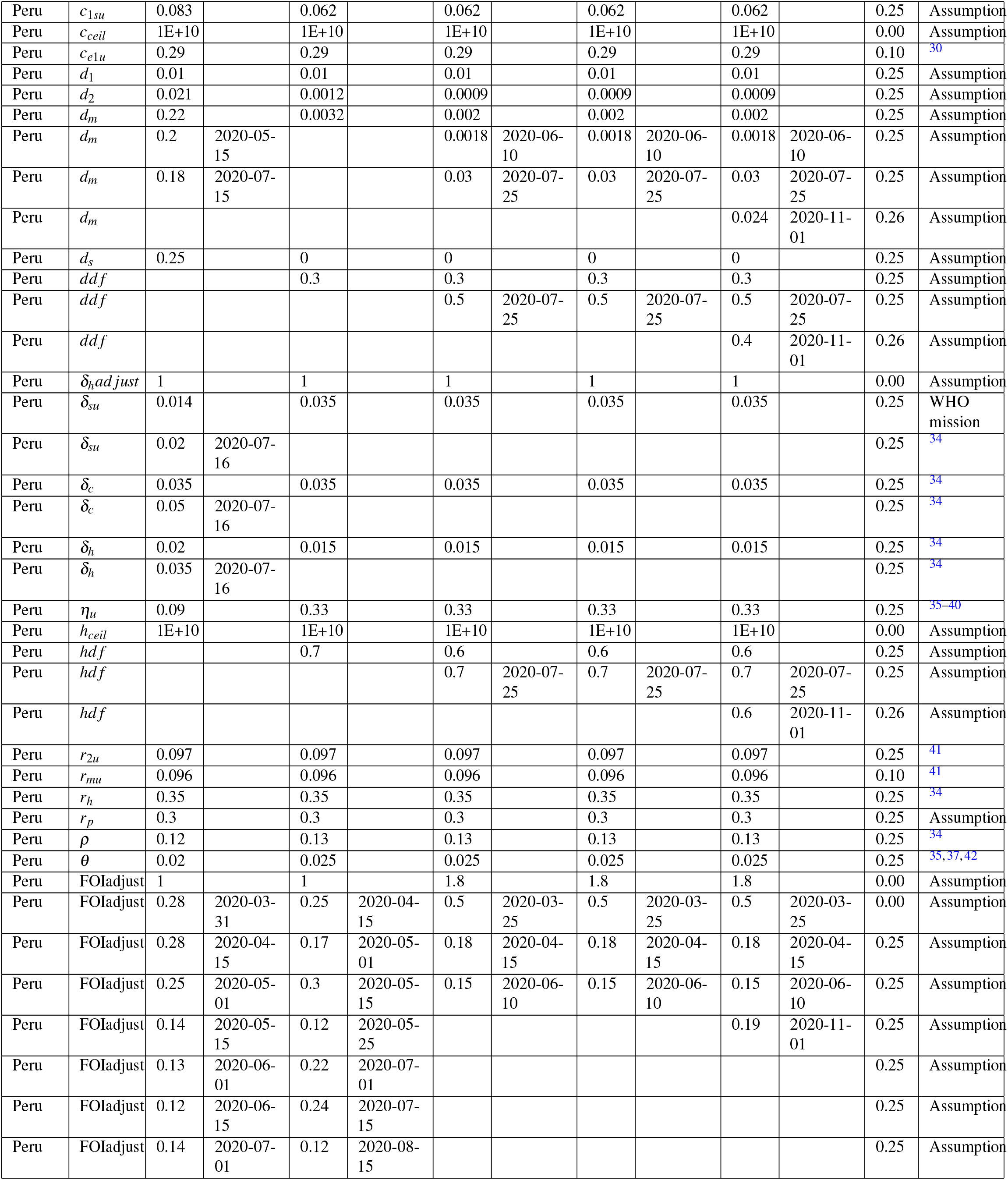
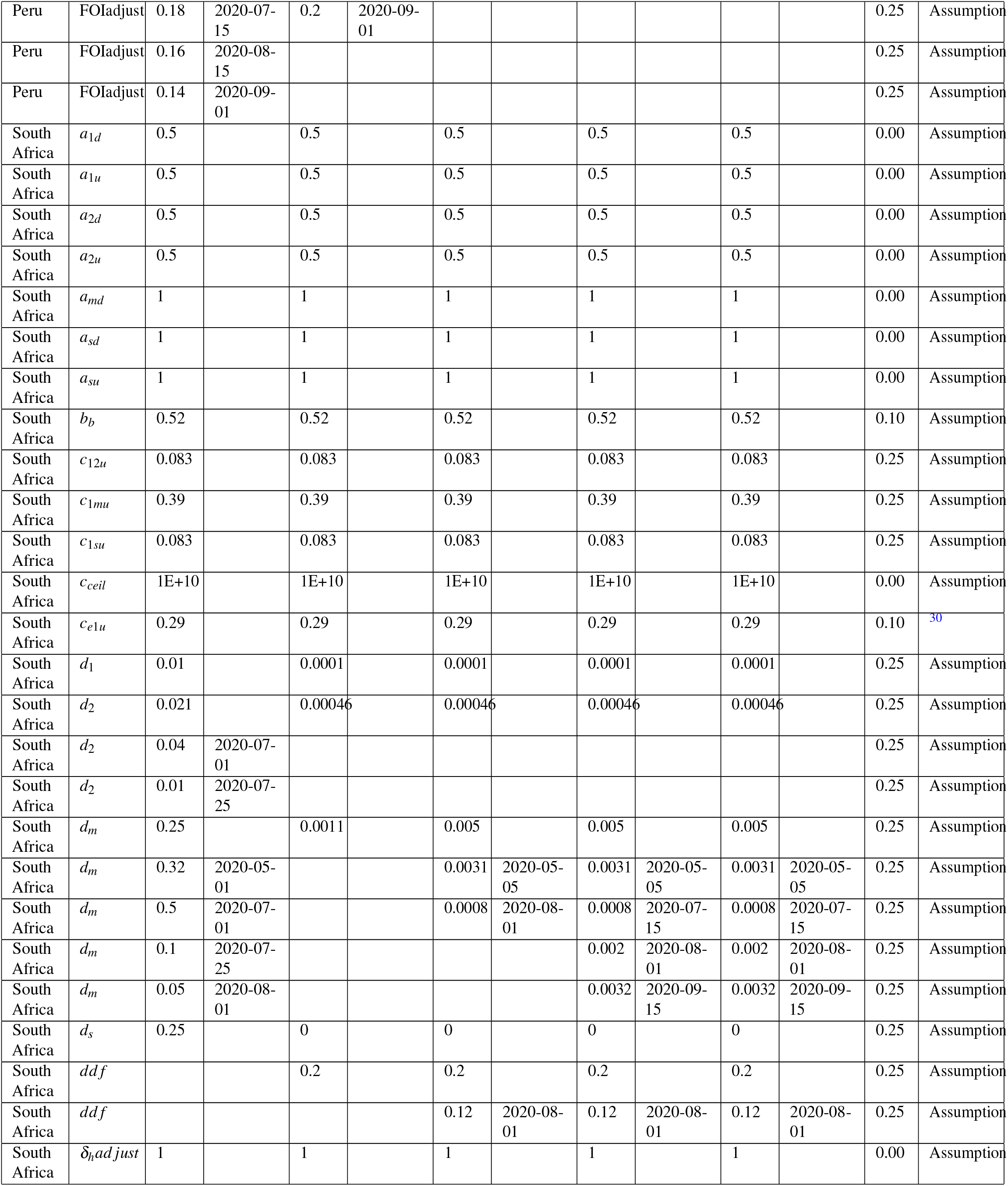
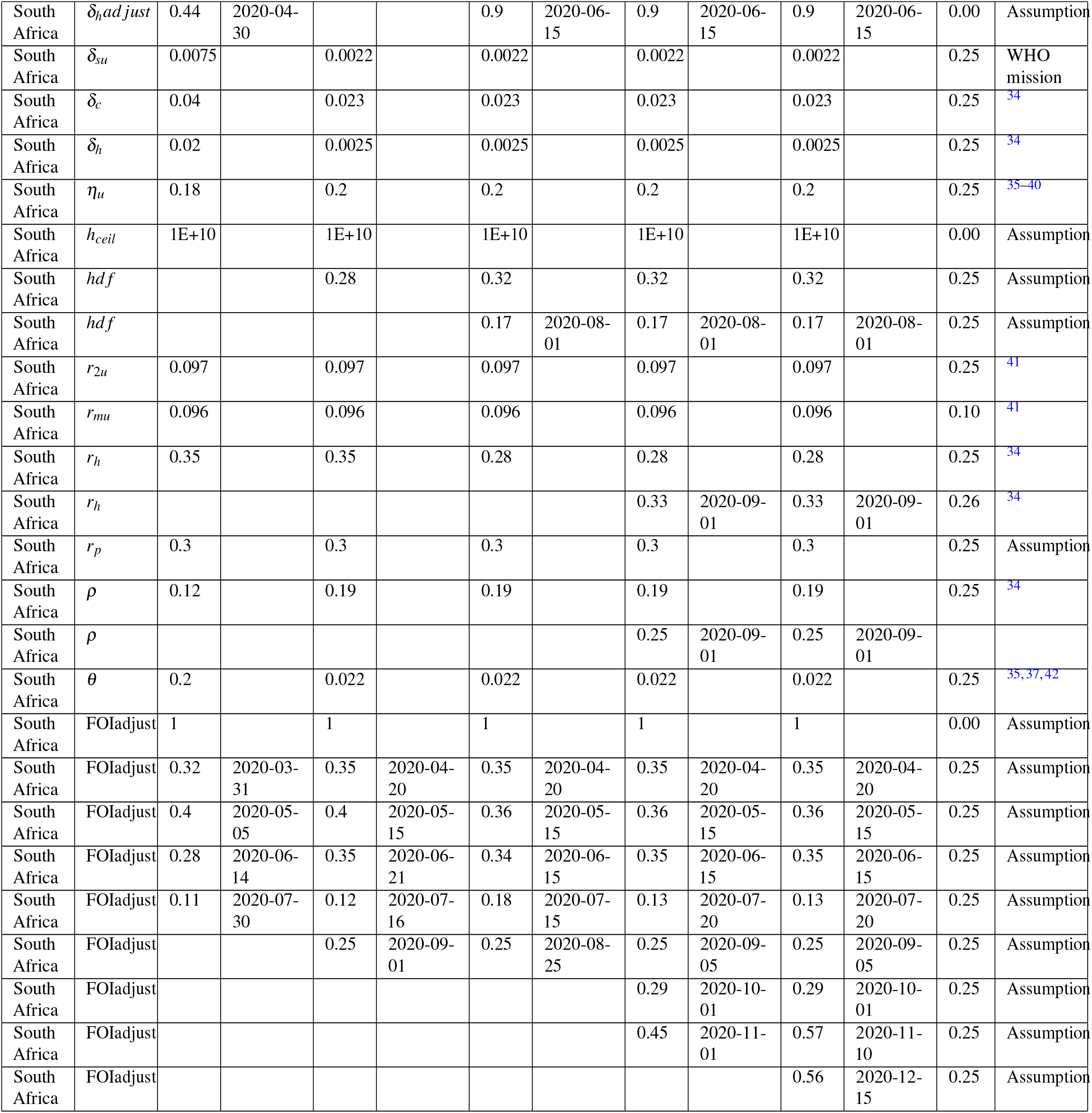
Model parameter values used in the model calibration for all the regions and calibration time points. Time-varying parameters have a corresponding start date. Delta column shows the range within which we allowed the parameters to vary. Fixed parameters have delta value equivalent to zero. Parameters that have blank values for some calibration time points indicate that those parameters were not part of the simulation model.For instance *dd f* and *hd f* were not parameters during model calibration for the first time point across all the regions.

| Region | Parameter | 2020/09/08 |  | 2020/10/13 |  | 2020/11/04 |  | 2020/12/14 |  | 2021/01/12 |  | Delta | Reference |
| --- | --- | --- | --- | --- | --- | --- | --- | --- | --- | --- | --- | --- | --- |
|  |  | Value | Start date | Value | Start date | Value | Start date | Value | Start date | Value | Start date |  |  |
| Brazil | $a_{1d}$ | 0.5 | | 0.5 | | 0.5 | | 0.5 | | 0.5 | | 0.00 | Assumption |
| Brazil | $a_{1u}$ | 0.5 | | 0.5 | | 0.5 | | 0.5 | | 0.5 | | 0.00 | Assumption |
| Brazil | $a_{2d}$ | 0.5 | | 0.5 | | 0.5 | | 0.5 | | 0.5 | | 0.00 | Assumption |
| Brazil | $a_{2u}$ | 0.5 | | 0.5 | | 0.5 | | 0.5 | | 0.5 | | 0.00 | Assumption |
| Brazil | $a_{md}$ | 1 | | 1 | | 1 | | 1 | | 1 | | 0.00 | Assumption |
| Brazil | $a_{sd}$ | 1 | | 1 | | 1 | | 1 | | 1 | | 0.00 | Assumption |
| Brazil | $a_{su}$ | 1 | | 1 | | 1 | | 1 | | 1 | | 0.00 | Assumption |
| Brazil | $b_b$ | 0.6 | | 0.6 | | 0.6 | | 0.6 | | 0.6 | | 0.10 | Assumption |
| Brazil | $c_{12u}$ | 0.083 | | 0.083 | | 0.083 | | 0.083 | | 0.083 | | 0.25 | Assumption |
| Brazil | $c_{1mu}$ | 0.39 | | 0.39 | | 0.39 | | 0.39 | | 0.39 | | 0.25 | Assumption |
| Brazil | $c_{1su}$ | 0.083 | | 0.083 | | 0.083 | | 0.083 | | 0.083 | | 0.25 | Assumption |
| Brazil | $c_{ceil}$ | 1E+10 | | 1E+10 | | 1E+10 | | 1E+10 | | 1E+10 | | 0.00 | Assumption |
| Brazil | $c_{elu}$ | 0.29 | | 0.29 | | 0.29 | | 0.29 | | 0.29 | | 0.10 | 30 |
| Brazil | $d_1$ | 0.01 | | 0 | | 0 | | 0 | | 0 | | 0.25 | Assumption |
| Brazil | $d_2$ | 0.021 | | 0.003 | | 0.003 | | 0.003 | | 0.003 | | 0.25 | Assumption |
| Brazil | $d_m$ | 0.21 | | 0.008 | | 0.0025 | | 0.0025 | | 0.0025 | | 0.25 | Assumption |
| Brazil | $d_m$ | 0.25 | 2020-04-15 | | | 0.008 | 2020-06-01 | 0.008 | 2020-06-01 | 0.008 | 2020-06-01 | 0.25 | Assumption |
| Brazil | $d_m$ | 0.25 | 2020-05-20 | | | 0.011 | 2020-07-20 | 0.011 | 2020-07-20 | 0.011 | 2020-07-20 | 0.25 | Assumption |
| Brazil | $d_m$ | | | | | | | | | 0.013 | 2020-11-15 | 0.25 | Assumption |
| Brazil | $d_s$ | 0.25 | | 0 | | 0 | | 0 | | 0 | | 0.25 | Assumption |
| Brazil | $ddf$ | | | 0.1 | | 0.1 | | 0.1 | | 0.1 | | 0.25 | Assumption |
| Brazil | $\delta_{hadjust}$ | 1 | | 1 | | 1 | | 1 | | 1 | | 0.00 | Assumption |
| Brazil | $\delta_{su}$ | 0.03 | | 0.0035 | | 0.0035 | | 0.0035 | | 0.0035 | | 0.25 | WHO mission |
| Brazil | $\delta_{su}$ | 0.021 | 2020-05-01 | | | | | | | | | 0.25 | WHO mission |
| Brazil | $\delta_{su}$ | 0.013 | 2020-07-15 | | | | | | | | | 0.25 | WHO mission |
| Brazil | $\delta_c$ | 0.04 | | 0.023 | | 0.023 | | 0.023 | | 0.023 | | 0.25 | 34 |
| Brazil | $\delta_c$ | 0.032 | 2020-05-01 | | | | | | | | | 0.25 | 34 |
| Brazil | $\delta_c$ | 0.017 | 2020-07-15 | | | | | | | | | 0.25 | 34 |
| Brazil | $\delta_h$ | 0.03 | | 0.0025 | | 0.0025 | | 0.0025 | | 0.0025 | | 0.25 | 34 |
| Brazil | $\delta_h$ | 0.023 | 2020-05-01 | | | | | | | | | 0.25 | 34 |
| Brazil | $\delta_h$ | 0.013 | 2020-07-15 | | | | | | | | | 0.25 | 34 |
| Brazil | $\eta_u$ | 0.15 | | 0.27 | | 0.27 | | 0.27 | | 0.27 | | 0.25 | 35–40 |
| Brazil | $h_{ceil}$ | 1E+10 | | 1E+10 | | 1E+10 | | 1E+10 | | 1E+10 | | 0.00 | Assumption |
| Brazil | $hdf$ | | | 0.25 | | 0.22 | | 0.22 | | 0.22 | | 0.25 | Assumption |
| Brazil | $hdf$ | | | | | 0.25 | 2020-06-01 | 0.25 | 2020-06-01 | 0.25 | 2020-06-01 | 0.25 | Assumption |
| Brazil | $hdf$ | | | | | | | 0.5 | 2020-10-15 | 0.5 | 2020-10-15 | 0.25 | Assumption |
| Brazil | $r_{2u}$ | 0.097 | | 0.097 | | 0.097 | | 0.097 | | 0.097 | | 0.25 | 41 |
| Brazil | $r_{mu}$ | 0.096 | | 0.096 | | 0.096 | | 0.096 | | 0.096 | | 0.10 | 41 |
| Brazil | $r_h$ | 0.35 | | 0.35 | | 0.35 | | 0.35 | | 0.35 | | 0.25 | 34 |
| Brazil | $r_p$ | 0.3 | | 0.3 | | 0.3 | | 0.3 | | 0.3 | | 0.25 | Assumption |
| Brazil | $\rho$ | 0.12 | | 0.19 | | 0.19 | | 0.19 | | 0.19 | | 0.25 | 34 |
| Brazil | $\theta$ | 0.02 | | 0.022 | | 0.022 | | 0.022 | | 0.022 | | 0.25 | 35,37,42 |
| Brazil | FOIadjust | 1 |  | 1.2 |  | 1.2 |  | 1.2 |  | 1.2 |  | 0.00 | Assumption |
| Brazil | FOIadjust | 0.61 | 2020-03-16 | 0.32 | 2020-04-05 | 0.3 | 2020-04-10 | 0.3 | 2020-04-10 | 0.3 | 2020-04-10 | 0.25 | Assumption |
| Brazil | FOIadjust | 0.36 | 2020-04-15 | 0.3 | 2020-05-01 | 0.26 | 2020-05-01 | 0.26 | 2020-05-01 | 0.26 | 2020-05-01 | 0.25 | Assumption |
| Brazil | FOIadjust | 0.18 | 2020-05-20 | 0.21 | 2020-06-01 | 0.18 | 2020-06-01 | 0.18 | 2020-06-01 | 0.18 | 2020-06-01 | 0.25 | Assumption |
| Brazil | FOIadjust |  |  | 0.24 | 2020-07-01 | 0.23 | 2020-07-01 | 0.23 | 2020-07-01 | 0.23 | 2020-07-01 | 0.25 | Assumption |
| Brazil | FOIadjust |  |  | 0.2 | 2020-08-01 | 0.21 | 2020-08-01 | 0.21 | 2020-08-01 | 0.21 | 2020-08-01 | 0.25 | Assumption |
| Brazil | FOIadjust |  |  |  |  |  |  | 0.29 | 2020-10-20 | 0.34 | 2020-11-01 | 0.25 | Assumption |
| Brazil | FOIadjust |  |  |  |  |  |  |  |  | 0.3 | 2020-12-01 | 0.25 | Assumption |
| Chile | $a_{1d}$ | 0.5 | | 0.5 | | 0.5 | | 0.5 | | 0.5 | | 0.00 | Assumption |
| Chile | $a_{1u}$ | 0.5 | | 0.5 | | 0.5 | | 0.5 | | 0.5 | | 0.00 | Assumption |
| Chile | $a_{2d}$ | 0.5 | | 0.5 | | 0.5 | | 0.5 | | 0.5 | | 0.00 | Assumption |
| Chile | $a_{2u}$ | 0.5 | | 0.5 | | 0.5 | | 0.5 | | 0.5 | | 0.00 | Assumption |
| Chile | $a_{md}$ | 1 | | 1 | | 1 | | 1 | | 1 | | 0.00 | Assumption |
| Chile | $a_{sd}$ | 1 | | 1 | | 1 | | 1 | | 1 | | 0.00 | Assumption |
| Chile | $a_{su}$ | 1 | | 1 | | 1 | | 1 | | 1 | | 0.00 | Assumption |
| Chile | $b_b$ | 0.49 | | 0.49 | | 0.49 | | 0.49 | | 0.49 | | 0.10 | Assumption |
| Chile | $c_{12u}$ | 0.083 | | 0.083 | | 0.083 | | 0.083 | | 0.083 | | 0.25 | Assumption |
| Chile | $c_{1mu}$ | 0.39 | | 0.39 | | 0.39 | | 0.39 | | 0.39 | | 0.25 | Assumption |
| Chile | $c_{1su}$ | 0.083 | | 0.083 | | 0.083 | | 0.083 | | 0.083 | | 0.25 | Assumption |
| Chile | $c_{ceil}$ | 1E+10 | | 1E+10 | | 1E+10 | | 1E+10 | | 1E+10 | | 0.00 | Assumption |
| Chile | $c_{e1u}$ | 0.29 | | 0.29 | | 0.29 | | 0.29 | | 0.29 | | 0.10 | 30 |
| Chile | $d_1$ | 0.01 | | 0.01 | | 0.01 | | 0.01 | | 0.01 | | 0.25 | Assumption |
| Chile | $d_2$ | 0.021 | | 0.021 | | 0.021 | | 0.021 | | 0.021 | | 0.25 | Assumption |
| Chile | $d_m$ | 0.25 | | 0.25 | | 0.25 | | 0.25 | | 0.25 | | 0.25 | Assumption |
| Chile | $d_m$ | 0.13 | 2020-06-01 | 0.12 | 2020-06-01 | 0.12 | 2020-06-01 | 0.12 | 2020-06-01 | 0.12 | 2020-06-01 | 0.25 | Assumption |
| Chile | $d_m$ | 0.07 | 2020-07-01 | 0.059 | 2020-07-01 | 0.06 | 2020-07-01 | 0.06 | 2020-07-01 | 0.06 | 2020-07-01 | 0.25 | Assumption |
| Chile | $d_m$ | | | 0.062 | 2020-08-15 | 0.07 | 2020-08-15 | 0.07 | 2020-08-15 | 0.07 | 2020-08-15 | 0.25 | Assumption |
| Chile | $d_m$ | | | | | | | | | 0.09 | 2020-12-01 | 0.25 | Assumption |
| Chile | $d_s$ | 0.25 | | 0.25 | | 0.25 | | 0.25 | | 0.25 | | 0.25 | Assumption |
| Chile | $ddf$ | | | 0.1 | | 0.1 | | 0.1 | | 0.1 | | 0.25 | Assumption |
| Chile | $ddf$ | | | | | | | | | 0.15 | 2020-12-01 | 0.25 | Assumption |
| Chile | $\delta_{hadjust}$ | 1 | | 1 | | 1 | | 1 | | 1 | | 0.00 | Assumption |
| Chile | $\delta_{su}$ | 0.009 | | 0.035 | | 0.035 | | 0.035 | | 0.035 | | 0.25 | WHO mission |
| Chile | $\delta_{su}$ | 0.0067 | 2020-04-20 | | | | | | | | | 0.25 | WHO mission |
| Chile | $\delta_c$ | 0.03 | | 0.035 | | 0.035 | | 0.035 | | 0.035 | | 0.25 | 34 |
| Chile | $\delta_c$ | 0.02 | 2020-04-20 | | | | | | | | | 0.25 | 34 |
| Chile | $\delta_h$ | 0.02 | | 0.015 | | 0.015 | | 0.015 | | 0.015 | | 0.25 | 34 |
| Chile | $\delta_h$ | 0.013 | 2020-04-20 | | | | | | | | | 0.25 | 34 |
| Chile | $\eta_u$ | 0.09 | | 0.33 | | 0.33 | | 0.33 | | 0.33 | | 0.25 | 35–40 |
| Chile | $\eta_u$ | | | | | | | | | 0.37 | 2020-12-01 | 0.25 | 35–40 |
| Chile | $h_{ceil}$ | 1E+10 | | 1E+10 | | 1E+10 | | 1E+10 | | 1E+10 | | 0.00 | Assumption |
| Chile | $hdf$ | | | 0.25 | | 0.25 | | 0.25 | | 0.25 | | 0.25 | Assumption |
| Chile | $hdf$ | | | | | | | | | 0.5 | 2020-12-01 | 0.25 | Assumption |
| Chile | $r_{2u}$ | 0.097 | | 0.097 | | 0.097 | | 0.097 | | 0.097 | | 0.25 | 41 |
| Chile | $r_{mu}$ | 0.096 | | 0.096 | | 0.096 | | 0.096 | | 0.096 | | 0.10 | 41 |
| Chile | $r_h$ | 0.35 | | 0.35 | | 0.35 | | 0.35 | | 0.35 | | 0.25 | 34 |
| Chile | $r_h$ | | | | | | | | | 0.37 | 2020-07-01 | 0.25 | 34 |
| Chile | $r_h$ | | | | | | | | | 0.4 | 2020-12-01 | 0.25 | 34 |
| Chile | $r_p$ | 0.3 | | 0.3 | | 0.3 | | 0.3 | | 0.3 | | 0.25 | Assumption |
| Chile | $\rho$ | 0.12 | | 0.13 | | 0.13 | | 0.13 | | 0.13 | | 0.25 | 34 |
| Chile | $\theta$ | 0.02 | | 0.025 | | 0.025 | | 0.025 | | 0.025 | | 0.25 | 35, 37, 42 |
| Chile | FOIadjust | 1 |  | 1.2 |  | 1.4 |  | 1.4 |  | 1.4 |  | 0.00 | Assumption |
| Chile | FOIadjust | 0.62 | 2020-03-23 | 0.41 | 2020-04-01 | 0.49 | 2020-03-25 | 0.49 | 2020-03-25 | 0.49 | 2020-03-25 | 0.25 | Assumption |
| Chile | FOIadjust | 0.48 | 2020-04-15 | 0.4 | 2020-05-12 | 0.44 | 2020-04-15 | 0.44 | 2020-04-15 | 0.44 | 2020-04-15 | 0.25 | Assumption |
| Chile | FOIadjust | 0.29 | 2020-05-15 | 0.19 | 2020-06-01 | 0.4 | 2020-05-14 | 0.4 | 2020-05-14 | 0.4 | 2020-05-14 | 0.25 | Assumption |
| Chile | FOIadjust | 0.18 | 2020-06-01 |  |  | 0.19 | 2020-06-01 | 0.19 | 2020-06-01 | 0.19 | 2020-06-01 | 0.25 | Assumption |
| Chile | FOIadjust | 0.16 | 2020-06-15 |  |  | 0.21 | 2020-08-15 | 0.21 | 2020-08-15 | 0.21 | 2020-08-15 | 0.25 | Assumption |
| Chile | FOIadjust |  |  |  |  |  |  | 0.23 | 2020-10-01 | 0.23 | 2020-10-01 | 0.25 | Assumption |
| Chile | FOIadjust |  |  |  |  |  |  | 0.21 | 2020-10-15 | 0.21 | 2020-10-15 | 0.25 | Assumption |
| Chile | FOIadjust |  |  |  |  |  |  |  |  | 0.25 | 2020-11-20 | 0.25 | Assumption |
| Limpopo | $a_{1d}$ | 0.5 | | 0.5 | | 0.5 | | 0.5 | | 0.5 | | 0.00 | Assumption |
| Limpopo | $a_{1u}$ | 0.5 | | 0.5 | | 0.5 | | 0.5 | | 0.5 | | 0.00 | Assumption |
| Limpopo | $a_{2d}$ | 0.5 | | 0.5 | | 0.5 | | 0.5 | | 0.5 | | 0.00 | Assumption |
| Limpopo | $a_{2u}$ | 0.5 | | 0.5 | | 0.5 | | 0.5 | | 0.5 | | 0.00 | Assumption |
| Limpopo | $a_{md}$ | 1 | | 1 | | 1 | | 1 | | 1 | | 0.00 | Assumption |
| Limpopo | $a_{sd}$ | 1 | | 1 | | 1 | | 1 | | 1 | | 0.00 | Assumption |
| Limpopo | $a_{su}$ | 1 | | 1 | | 1 | | 1 | | 1 | | 0.00 | Assumption |
| Limpopo | $b_b$ | 0.25 | | 0.25 | | 0.25 | | 0.25 | | 0.25 | | 0.10 | Assumption |
| Limpopo | $c_{12u}$ | 0.083 | | 0.083 | | 0.083 | | 0.083 | | 0.083 | | 0.25 | Assumption |
| Limpopo | $c_{1mu}$ | 0.39 | | 0.39 | | 0.39 | | 0.39 | | 0.39 | | 0.25 | Assumption |
| Limpopo | $c_{1su}$ | 0.083 | | 0.083 | | 0.083 | | 0.083 | | 0.083 | | 0.25 | Assumption |
| Limpopo | $c_{ceil}$ | 1E+10 | | 1E+10 | | 1E+10 | | 1E+10 | | 1E+10 | | 0.00 | Assumption |
| Limpopo | $c_{e1u}$ | 0.29 | | 0.29 | | 0.29 | | 0.29 | | 0.29 | | 0.10 | 30 |
| Limpopo | $d_1$ | 0.01 | | 0 | | 0 | | 0 | | 0 | | 0.25 | Assumption |
| Limpopo | $d_2$ | 0.021 | | 0.0003 | | 0.0003 | | 0.0003 | | 0.0003 | | 0.25 | Assumption |
| Limpopo | $d_m$ | 0.21 | | 0.0008 | | 0.0008 | | 0.0008 | | 0.0008 | | 0.25 | Assumption |
| Limpopo | $d_m$ | 0.16 | 2020-05-01 | | | | | | | | | 0.25 | Assumption |
| Limpopo | $d_m$ | 0.15 | 2020-08-15 | | | | | | | | | 0.25 | Assumption |
| Limpopo | $d_s$ | 0.25 | | 0 | | 0 | | 0 | | 0 | | 0.25 | Assumption |
| Limpopo | $ddf$ | | | 0.1 | | 0.1 | | 0.1 | | 0.1 | | 0.25 | Assumption |
| Limpopo | $\delta_{hadjust}$ | 1 | | 0.92 | | 0.92 | | 0.92 | | 0.92 | | 0.00 | Assumption |
| Limpopo | $\delta_{hadjust}$ | 0.38 | 2020-05-15 | | | | | | | | | 0.25 | Assumption |
| Limpopo | $\delta_{su}$ | 0.004 | | 0.0035 | | 0.0035 | | 0.0035 | | 0.0035 | | 0.25 | WHO mission |
| Limpopo | $\delta_c$ | 0.04 | | 0.023 | | 0.023 | | 0.023 | | 0.023 | | 0.25 | 34 |
| Limpopo | $\delta_h$ | 0.03 | | 0.0025 | | 0.0025 | | 0.0025 | | 0.0025 | | 0.25 | 34 |
| Limpopo | $\eta_u$ | 0.18 | | 0.25 | | 0.25 | | 0.25 | | 0.25 | | 0.25 | 35–40 |
| Limpopo | $h_{ceil}$ | 1E+10 | | 1E+10 | | 1E+10 | | 1E+10 | | 1E+10 | | 0.00 | Assumption |
| Limpopo | $hdf$ | | | 0.2 | | 0.2 | | 0.2 | | 0.2 | | 0.25 | Assumption |
| Limpopo | $r_{2u}$ | 0.097 | | 0.097 | | 0.097 | | 0.097 | | 0.097 | | 0.25 | 41 |
| Limpopo | $r_{mu}$ | 0.096 | | 0.096 | | 0.096 | | 0.096 | | 0.096 | | 0.10 | 41 |
| Limpopo | $r_h$ | 0.35 | | 0.35 | | 0.35 | | 0.35 | | 0.35 | | 0.25 | 34 |
| Limpopo | $r_p$ | 0.3 | | 0.3 | | 0.3 | | 0.3 | | 0.3 | | 0.25 | Assumption |
| Limpopo | $\rho$ | 0.12 | | 0.19 | | 0.19 | | 0.19 | | 0.19 | | 0.25 | 34 |
| Limpopo | $\theta$ | 0.2 | | 0.022 | | 0.022 | | 0.022 | | 0.022 | | 0.25 | 35,37,42 |
| Limpopo | FOIadjust | 1 |  | 1.3 |  | 1.3 |  | 1.3 |  | 1.3 |  | 0.00 | Assumption |
| Limpopo | FOIadjust | 0.4 | 2020-04-01 | 1 | 2020-04-11 | 1 | 2020-04-11 | 1.3 |  | 1.3 |  | 0.25 | Assumption |
| Limpopo | FOIadjust | 0.77 | 2020-04-20 | 0.82 | 2020-04-20 | 0.82 | 2020-04-20 | 1 | 2020-04-11 | 1 | 2020-04-11 | 0.25 | Assumption |
| Limpopo | FOIadjust | 0.8 | 2020-06-15 | 1 | 2020-06-15 | 1 | 2020-06-15 | 0.82 | 2020-04-20 | 0.82 | 2020-04-20 | 0.25 | Assumption |
| Limpopo | FOIadjust | 0.7 | 2020-07-01 | 0.8 | 2020-07-01 | 0.8 | 2020-07-01 | 1 | 2020-06-15 | 1 | 2020-06-15 | 0.25 | Assumption |
| Limpopo | FOIadjust | 0.5 | 2020-07-15 | 0.48 | 2020-07-15 | 0.48 | 2020-07-15 | 0.8 | 2020-07-01 | 0.8 | 2020-07-01 | 0.25 | Assumption |
| Limpopo | FOIadjust | 0.24 | 2020-08-01 | 0.21 | 2020-08-01 | 0.21 | 2020-08-01 | 0.48 | 2020-07-15 | 0.48 | 2020-07-15 | 0.25 | Assumption |
| Limpopo | FOIadjust | 0.2 | 2020-08-15 | 0.45 | 2020-09-01 | 0.45 | 2020-09-01 | 0.21 | 2020-08-01 | 0.21 | 2020-08-01 | 0.25 | Assumption |
| Limpopo | FOIadjust |  |  |  |  |  |  | 0.45 | 2020-09-01 | 0.45 | 2020-09-01 | 0.25 | Assumption |
| Limpopo | FOIadjust |  |  |  |  |  |  | 0.35 | 2020-10-11 | 0.35 | 2020-10-11 | 0.25 | Assumption |
| Limpopo | FOIadjust |  |  |  |  |  |  |  |  | 1 | 2020-11-15 | 0.25 | Assumption |
| Limpopo | FOIadjust |  |  |  |  |  |  |  |  | 1 | 2020-12-15 | 0.25 | Assumption |
| North West | $a_{1d}$ | 0.5 | | 0.5 | | 0.5 | | 0.5 | | 0.5 | | 0.00 | Assumption |
| North West | $a_{1u}$ | 0.5 | | 0.5 | | 0.5 | | 0.5 | | 0.5 | | 0.00 | Assumption |
| North West | $a_{2d}$ | 0.5 | | 0.5 | | 0.5 | | 0.5 | | 0.5 | | 0.00 | Assumption |
| North West | $a_{2u}$ | 0.5 | | 0.5 | | 0.5 | | 0.5 | | 0.5 | | 0.00 | Assumption |
| North West | $a_{md}$ | 1 | | 1 | | 1 | | 1 | | 1 | | 0.00 | Assumption |
| North West | $a_{sd}$ | 1 | | 1 | | 1 | | 1 | | 1 | | 0.00 | Assumption |
| North West | $a_{su}$ | 1 | | 1 | | 1 | | 1 | | 1 | | 0.00 | Assumption |
| North West | $b_b$ | 0.22 | | 0.22 | | 0.22 | | 0.22 | | 0.22 | | 0.10 | Assumption |
| North West | $c_{12u}$ | 0.083 | | 0.083 | | 0.083 | | 0.083 | | 0.083 | | 0.25 | Assumption |
| North West | $c_{1mu}$ | 0.39 | | 0.39 | | 0.39 | | 0.39 | | 0.39 | | 0.25 | Assumption |
| North West | $c_{1su}$ | 0.083 | | 0.083 | | 0.083 | | 0.083 | | 0.083 | | 0.25 | Assumption |
| North West | $c_{ceil}$ | 1E+10 | | 1E+10 | | 1E+10 | | 1E+10 | | 1E+10 | | 0.00 | Assumption |
| North West | $c_{elu}$ | 0.29 | | 0.29 | | 0.29 | | 0.29 | | 0.29 | | 0.10 | 30 |
| North West | $d_1$ | 0.01 | | 0 | | 0 | | 0 | | 0 | | 0.25 | Assumption |
| North West | $d_2$ | 0.021 | | 0.0005 | | 0.0005 | | 0.0005 | | 0.0005 | | 0.25 | Assumption |
| North West | $d_m$ | 0.21 | | 0.002 | | 0.002 | | 0.002 | | 0.002 | | 0.25 | Assumption |
| North West | $d_m$ | 0.21 | 2020-05-15 | | | | | | | | | 0.25 | Assumption |
| North West | $d_m$ | 0.3 | 2020-06-01 | | | | | | | | | 0.25 | Assumption |
| North West | $d_m$ | 0.08 | 2020-08-01 | | | | | | | | | 0.25 | Assumption |
| North West | $d_s$ | 0.25 | | 0 | | 0 | | 0 | | 0 | | 0.25 | Assumption |
| North West | $ddf$ | | | 0.13 | | 0.13 | | 0.13 | | 0.13 | | 0.25 | Assumption |
| North West | $\delta_{hadjust}$ | 1 | | 1 | | 1 | | 1 | | 1 | | 0.00 | Assumption |
| North West | $\delta_{hadjust}$ | 0.3 | 2020-05-15 | | | | | | | | | 0.25 | Assumption |
| North West | $\delta_{su}$ | 0.004 | | 0.0035 | | 0.0035 | | 0.0035 | | 0.0035 | | 0.25 | WHO mission |
| North West | $\delta_c$ | 0.021 | | 0.023 | | 0.023 | | 0.023 | | 0.023 | | 0.25 | 34 |
| North West | $\delta_h$ | 0.013 | | 0.0025 | | 0.0025 | | 0.0025 | | 0.0025 | | 0.25 | 34 |
| North West | $\delta_h$ | 0.013 | 2020-07-01 | | | | | | | | | 0.25 | 34 |
| North West | $\delta_h$ | 0.021 | 2020-08-01 | | | | | | | | | | 34 |
| North West | $\eta_u$ | 0.18 | | 0.25 | | 0.25 | | 0.25 | | 0.25 | | | 35–40 |
| North West | $h_{ceil}$ | 1E+10 | | 1E+10 | | 1E+10 | | 1E+10 | | 1E+10 | | 0.00 | Assumption |
| North West | $hdf$ | | | 0.25 | | 0.25 | | 0.25 | | 0.25 | | 0.25 | Assumption |
| North West | $r_{2u}$ | 0.097 | | 0.097 | | 0.097 | | 0.097 | | 0.097 | | 0.25 | 41 |
| North West | $r_{mu}$ | 0.096 | | 0.096 | | 0.096 | | 0.096 | | 0.096 | | 0.10 | 41 |
| North West | $r_h$ | 0.35 | | 0.35 | | 0.35 | | 0.35 | | 0.35 | | 0.25 | 34 |
| North West | $r_p$ | 0.3 | | 0.3 | | 0.3 | | 0.3 | | 0.3 | | 0.25 | Assumption |
| North West | $\rho$ | 0.12 | | 0.19 | | 0.19 | | 0.19 | | 0.19 | | 0.25 | 34 |
| North West | $\theta$ | 0.2 | | 0.022 | | 0.022 | | 0.022 | | 0.022 | | 0.25 | 35,37,42 |
| North West | FOIadjust | 1 |  | 1.3 |  | 1.3 |  | 1.3 |  | 1.3 |  | 0.00 | Assumption |
| North West | FOIadjust | 0.88 | 2020-05-10 | 0.95 | 2020-06-25 | 0.95 | 2020-06-25 | 0.95 | 2020-06-25 | 0.95 | 2020-06-25 | 0.25 | Assumption |
| North West | FOIadjust | 0.82 | 2020-06-13 | 0.35 | 2020-07-08 | 0.35 | 2020-07-08 | 0.35 | 2020-07-08 | 0.35 | 2020-07-08 | 0.25 | Assumption |
| North West | FOIadjust | 0.56 | 2020-07-20 | 0.27 | 2020-08-01 | 0.27 | 2020-08-01 | 0.27 | 2020-08-01 | 0.27 | 2020-08-01 | 0.25 | Assumption |
| North West | FOIadjust | 0.25 | 2020-08-01 | 0.53 | 2020-08-18 | 0.58 | 2020-08-21 | 0.58 | 2020-08-21 | 0.58 | 2020-08-21 | 0.25 | Assumption |
| North West | FOIadjust |  |  |  |  |  |  | 0.4 | 2020-10-10 | 0.4 | 2020-10-10 | 0.25 | Assumption |
| North West | FOIadjust |  |  |  |  |  |  |  |  | 1.1 | 2020-11-15 | 0.25 | Assumption |
| Northern Cape | $a_{1d}$ | 0.5 | | 0.5 | | 0.5 | | 0.5 | | 0.5 | | 0.00 | Assumption |
| Northern Cape | $a_{1u}$ | 0.5 | | 0.5 | | 0.5 | | 0.5 | | 0.5 | | 0.00 | Assumption |
| Northern Cape | $a_{2d}$ | 0.5 | | 0.5 | | 0.5 | | 0.5 | | 0.5 | | 0.00 | Assumption |
| Northern Cape | $a_{2u}$ | 0.5 | | 0.5 | | 0.5 | | 0.5 | | 0.5 | | 0.00 | Assumption |
| Northern Cape | $a_{md}$ | 1 | | 1 | | 1 | | 1 | | 1 | | 0.00 | Assumption |
| Northern Cape | $a_{sd}$ | 1 | | 1 | | 1 | | 1 | | 1 | | 0.00 | Assumption |
| Northern Cape | $a_{su}$ | 1 | | 1 | | 1 | | 1 | | 1 | | 0.00 | Assumption |
| Northern Cape | $b_b$ | 0.24 | | 0.24 | | 0.24 | | 0.24 | | 0.24 | | 0.10 | Assumption |
| Northern Cape | $c_{12u}$ | 0.083 | | 0.083 | | 0.083 | | 0.083 | | 0.083 | | 0.25 | Assumption |
| Northern Cape | $c_{1mu}$ | 0.39 | | 0.39 | | 0.39 | | 0.39 | | 0.39 | | 0.25 | Assumption |
| Northern Cape | $c_{1su}$ | 0.083 | | 0.083 | | 0.083 | | 0.083 | | 0.083 | | 0.25 | Assumption |
| Northern Cape | $c_{ceil}$ | 1E+10 | | 1E+10 | | 1E+10 | | 1E+10 | | 1E+10 | | 0.00 | Assumption |
| Northern Cape | $c_{e1u}$ | 0.29 | | 0.29 | | 0.29 | | 0.29 | | 0.29 | | 0.10 | 30 |
| Northern Cape | $d_1$ | 0.01 | | 0 | | 0 | | 0 | | 0 | | 0.25 | Assumption |
| Northern Cape | $d_2$ | 0.021 | | 0.0002 | | 0.0002 | | 0.0002 | | 0.0002 | | 0.25 | Assumption |
| Northern Cape | $d_2$ | 0.1 | 2020-07-01 | | | | | | | | | 0.25 | Assumption |
| Northern Cape | $d_m$ | 0.21 | | 0.0008 | | 0.0008 | | 0.0008 | | 0.0008 | | 0.25 | Assumption |
| Northern Cape | $d_m$ | 0.5 | 2020-07-01 | | | | | 0.0015 | 2020-08-01 | 0.0015 | 2020-08-01 | 0.25 | Assumption |
| Northern Cape | $d_m$ | 0.3 | 2020-08-01 | | | | | | | | | 0.25 | Assumption |
| Northern Cape | $d_s$ | 0.25 | | 0 | | 0 | | 0 | | 0 | | 0.25 | Assumption |
| Northern Cape | $ddf$ | | | 0.2 | | 0.2 | | 0.2 | | 0.2 | | 0.25 | Assumption |
| Northern Cape | $\delta_{hadjust}$ | 1 | | 1 | | 1 | | 1 | | 1 | | 0.00 | Assumption |
| Northern Cape | $\delta_{hadjust}$ | 0.55 | 2020-05-22 | | | | | | | | | 0.25 | Assumption |
| Northern Cape | $\delta_{su}$ | 0.004 | | 0.0035 | | 0.0035 | | 0.0035 | | 0.0035 | | 0.25 | WHO mission |
| Northern Cape | $\delta_c$ | 0.02 | | 0.023 | | 0.023 | | 0.027 | | 0.027 | | 0.25 | 34 |
| Northern Cape | $\delta_h$ | 0.02 | | 0.0025 | | 0.0025 | | 0.003 | | 0.0035 | | 0.25 | 34 |
| Northern Cape | $\delta_h$ | | | | | | | | | 0.003 | 2020-09-01 | 0.25 | 34 |
| Northern Cape | $\delta_h$ | | | | | | | | | 0.0035 | 2020-11-15 | 0.25 | 34 |
| Northern Cape | $\eta_u$ | 0.18 | | 0.25 | | 0.25 | | 0.25 | | 0.25 | | 0.25 | 35–40 |
| Northern Cape | $\eta_u$ | | | | | | | | | 0.2 | 2020-11-15 | 0.25 | 35–40 |
| Northern Cape | $h_{ceil}$ | 1E+10 | | 1E+10 | | 1E+10 | | 1E+10 | | 1E+10 | | 0.00 | Assumption |
| Northern Cape | $hdf$ | | | 0.35 | | 0.35 | | 0.35 | | 0.35 | | 0.25 | Assumption |
| Northern Cape | $r_{2u}$ | 0.097 | | 0.097 | | 0.097 | | 0.097 | | 0.097 | | 0.25 | 41 |
| Northern Cape | $r_{mu}$ | 0.096 | | 0.096 | | 0.096 | | 0.096 | | 0.096 | | 0.10 | 41 |
| Northern Cape | $r_h$ | 0.35 | | 0.35 | | 0.35 | | 0.35 | | 0.32 | | 0.25 | 34 |
| Northern Cape | $r_h$ | | | | | | | | | 0.44 | 2020-09-01 | 0.25 | 34 |
| Northern Cape | $r_p$ | 0.3 | | 0.3 | | 0.3 | | 0.3 | | 0.3 | | 0.25 | Assumption |
| Northern Cape | $\rho$ | 0.12 | | 0.19 | | 0.19 | | 0.19 | | 0.19 | | 0.25 | 34 |
| Northern Cape | $\theta$ | 0.2 | | 0.022 | | 0.022 | | 0.022 | | 0.022 | | 0.25 | 35, 37, 42 |
| Northern Cape | FOIadjust | 1 |  | 1 |  | 1 |  | 1 |  | 1 |  | 0.25 | Assumption |
| Northern Cape | FOIadjust | 0.15 | 2020-04-02 | 0.85 | 2020-05-15 | 0.85 | 2020-05-15 | 0.85 | 2020-05-15 | 0.85 | 2020-05-15 | 0.25 | Assumption |
| Northern Cape | FOIadjust | 0.95 | 2020-04-27 | 1 | 2020-05-28 | 1 | 2020-05-28 | 1 | 2020-05-28 | 1 | 2020-05-28 | 0.25 | Assumption |
| Northern Cape | FOIadjust | 0.5 | 2020-07-20 | 0.59 | 2020-07-12 | 0.59 | 2020-07-12 | 0.58 | 2020-07-12 | 0.58 | 2020-07-12 | 0.25 | Assumption |
| Northern Cape | FOIadjust |  |  |  |  |  |  | 0.4 | 2020-10-10 | 0.44 | 2020-10-10 | 0.25 | Assumption |
| Northern Cape | FOIadjust |  |  |  |  |  |  |  |  | 1.2 | 2020-11-25 | 0.25 | Assumption |
| Northern Cape | FOIadjust |  |  |  |  |  |  |  |  | 1.4 | 2020-12-21 | 0.25 | Assumption |
| Peru | $a_{1d}$ | 0.5 | | 0.5 | | 0.5 | | 0.5 | | 0.5 | | 0.00 | Assumption |
| Peru | $a_{1u}$ | 0.5 | | 0.5 | | 0.5 | | 0.5 | | 0.5 | | 0.00 | Assumption |
| Peru | $a_{2d}$ | 0.5 | | 0.5 | | 0.5 | | 0.5 | | 0.5 | | 0.00 | Assumption |
| Peru | $a_{2u}$ | 0.5 | | 0.5 | | 0.5 | | 0.5 | | 0.5 | | 0.00 | Assumption |
| Peru | $a_{md}$ | 1 | | 1 | | 1 | | 1 | | 1 | | 0.00 | Assumption |
| Peru | $a_{sd}$ | 1 | | 1 | | 1 | | 1 | | 1 | | 0.00 | Assumption |
| Peru | $a_{su}$ | 1 | | 1 | | 1 | | 1 | | 1 | | 0.00 | Assumption |
| Peru | $b_b$ | 0.71 | | 0.71 | | 0.71 | | 0.71 | | 0.71 | | 0.10 | Assumption |
| Peru | $c_{12u}$ | 0.083 | | 0.092 | | 0.092 | | 0.092 | | 0.092 | | 0.25 | Assumption |
| Peru | $c_{1mu}$ | 0.39 | | 0.4 | | 0.4 | | 0.4 | | 0.4 | | 0.25 | Assumption |
| Peru | $c_{1su}$ | 0.083 | | 0.062 | | 0.062 | | 0.062 | | 0.062 | | 0.25 | Assumption |
| Peru | $c_{ceil}$ | 1E+10 | | 1E+10 | | 1E+10 | | 1E+10 | | 1E+10 | | 0.00 | Assumption |
| Peru | $c_{e1u}$ | 0.29 | | 0.29 | | 0.29 | | 0.29 | | 0.29 | | 0.10 | 30 |
| Peru | $d_1$ | 0.01 | | 0.01 | | 0.01 | | 0.01 | | 0.01 | | 0.25 | Assumption |
| Peru | $d_2$ | 0.021 | | 0.0012 | | 0.0009 | | 0.0009 | | 0.0009 | | 0.25 | Assumption |
| Peru | $d_m$ | 0.22 | | 0.0032 | | 0.002 | | 0.002 | | 0.002 | | 0.25 | Assumption |
| Peru | $d_m$ | 0.2 | 2020-05-15 | | | 0.0018 | 2020-06-10 | 0.0018 | 2020-06-10 | 0.0018 | 2020-06-10 | 0.25 | Assumption |
| Peru | $d_m$ | 0.18 | 2020-07-15 | | | 0.03 | 2020-07-25 | 0.03 | 2020-07-25 | 0.03 | 2020-07-25 | 0.25 | Assumption |
| Peru | $d_m$ | | | | | | | | | 0.024 | 2020-11-01 | 0.26 | Assumption |
| Peru | $d_s$ | 0.25 | | 0 | | 0 | | 0 | | 0 | | 0.25 | Assumption |
| Peru | $ddf$ | | | 0.3 | | 0.3 | | 0.3 | | 0.3 | | 0.25 | Assumption |
| Peru | $ddf$ | | | | | 0.5 | 2020-07-25 | 0.5 | 2020-07-25 | 0.5 | 2020-07-25 | 0.25 | Assumption |
| Peru | $ddf$ | | | | | | | | | 0.4 | 2020-11-01 | 0.26 | Assumption |
| Peru | $\delta_{hadjust}$ | 1 | | 1 | | 1 | | 1 | | 1 | | 0.00 | Assumption |
| Peru | $\delta_{su}$ | 0.014 | | 0.035 | | 0.035 | | 0.035 | | 0.035 | | 0.25 | WHO mission |
| Peru | $\delta_{su}$ | 0.02 | 2020-07-16 | | | | | | | | | 0.25 | 34 |
| Peru | $\delta_c$ | 0.035 | | 0.035 | | 0.035 | | 0.035 | | 0.035 | | 0.25 | 34 |
| Peru | $\delta_c$ | 0.05 | 2020-07-16 | | | | | | | | | 0.25 | 34 |
| Peru | $\delta_h$ | 0.02 | | 0.015 | | 0.015 | | 0.015 | | 0.015 | | 0.25 | 34 |
| Peru | $\delta_h$ | 0.035 | 2020-07-16 | | | | | | | | | 0.25 | 34 |
| Peru | $\eta_u$ | 0.09 | | 0.33 | | 0.33 | | 0.33 | | 0.33 | | 0.25 | 35–40 |
| Peru | $h_{ceil}$ | 1E+10 | | 1E+10 | | 1E+10 | | 1E+10 | | 1E+10 | | 0.00 | Assumption |
| Peru | $hdf$ | | | 0.7 | | 0.6 | | 0.6 | | 0.6 | | 0.25 | Assumption |
| Peru | $hdf$ | | | | | 0.7 | 2020-07-25 | 0.7 | 2020-07-25 | 0.7 | 2020-07-25 | 0.25 | Assumption |
| Peru | $hdf$ | | | | | | | | | 0.6 | 2020-11-01 | 0.26 | Assumption |
| Peru | $r_{2u}$ | 0.097 | | 0.097 | | 0.097 | | 0.097 | | 0.097 | | 0.25 | 41 |
| Peru | $r_{mu}$ | 0.096 | | 0.096 | | 0.096 | | 0.096 | | 0.096 | | 0.10 | 41 |
| Peru | $r_h$ | 0.35 | | 0.35 | | 0.35 | | 0.35 | | 0.35 | | 0.25 | 34 |
| Peru | $r_p$ | 0.3 | | 0.3 | | 0.3 | | 0.3 | | 0.3 | | 0.25 | Assumption |
| Peru | $\rho$ | 0.12 | | 0.13 | | 0.13 | | 0.13 | | 0.13 | | 0.25 | 34 |
| Peru | $\theta$ | 0.02 | | 0.025 | | 0.025 | | 0.025 | | 0.025 | | 0.25 | 35,37,42 |
| Peru | FOIadjust | 1 |  | 1 |  | 1.8 |  | 1.8 |  | 1.8 |  | 0.00 | Assumption |
| Peru | FOIadjust | 0.28 | 2020-03-31 | 0.25 | 2020-04-15 | 0.5 | 2020-03-25 | 0.5 | 2020-03-25 | 0.5 | 2020-03-25 | 0.00 | Assumption |
| Peru | FOIadjust | 0.28 | 2020-04-15 | 0.17 | 2020-05-01 | 0.18 | 2020-04-15 | 0.18 | 2020-04-15 | 0.18 | 2020-04-15 | 0.25 | Assumption |
| Peru | FOIadjust | 0.25 | 2020-05-01 | 0.3 | 2020-05-15 | 0.15 | 2020-06-10 | 0.15 | 2020-06-10 | 0.15 | 2020-06-10 | 0.25 | Assumption |
| Peru | FOIadjust | 0.14 | 2020-05-15 | 0.12 | 2020-05-25 |  |  |  |  | 0.19 | 2020-11-01 | 0.25 | Assumption |
| Peru | FOIadjust | 0.13 | 2020-06-01 | 0.22 | 2020-07-01 |  |  |  |  |  |  | 0.25 | Assumption |
| Peru | FOIadjust | 0.12 | 2020-06-15 | 0.24 | 2020-07-15 |  |  |  |  |  |  | 0.25 | Assumption |
| Peru | FOIadjust | 0.14 | 2020-07-01 | 0.12 | 2020-08-15 |  |  |  |  |  |  | 0.25 | Assumption |
| Peru | FOIadjust | 0.18 | 2020-07-15 | 0.2 | 2020-09-01 |  |  |  |  |  |  | 0.25 | Assumption |
| Peru | FOIadjust | 0.16 | 2020-08-15 |  |  |  |  |  |  |  |  | 0.25 | Assumption |
| Peru | FOIadjust | 0.14 | 2020-09-01 |  |  |  |  |  |  |  |  | 0.25 | Assumption |
| South Africa | $a_{1d}$ | 0.5 | | 0.5 | | 0.5 | | 0.5 | | 0.5 | | 0.00 | Assumption |
| South Africa | $a_{1u}$ | 0.5 | | 0.5 | | 0.5 | | 0.5 | | 0.5 | | 0.00 | Assumption |
| South Africa | $a_{2d}$ | 0.5 | | 0.5 | | 0.5 | | 0.5 | | 0.5 | | 0.00 | Assumption |
| South Africa | $a_{2u}$ | 0.5 | | 0.5 | | 0.5 | | 0.5 | | 0.5 | | 0.00 | Assumption |
| South Africa | $a_{md}$ | 1 | | 1 | | 1 | | 1 | | 1 | | 0.00 | Assumption |
| South Africa | $a_{sd}$ | 1 | | 1 | | 1 | | 1 | | 1 | | 0.00 | Assumption |
| South Africa | $a_{su}$ | 1 | | 1 | | 1 | | 1 | | 1 | | 0.00 | Assumption |
| South Africa | $b_b$ | 0.52 | | 0.52 | | 0.52 | | 0.52 | | 0.52 | | 0.10 | Assumption |
| South Africa | $c_{12u}$ | 0.083 | | 0.083 | | 0.083 | | 0.083 | | 0.083 | | 0.25 | Assumption |
| South Africa | $c_{1mu}$ | 0.39 | | 0.39 | | 0.39 | | 0.39 | | 0.39 | | 0.25 | Assumption |
| South Africa | $c_{1su}$ | 0.083 | | 0.083 | | 0.083 | | 0.083 | | 0.083 | | 0.25 | Assumption |
| South Africa | $c_{ceil}$ | 1E+10 | | 1E+10 | | 1E+10 | | 1E+10 | | 1E+10 | | 0.00 | Assumption |
| South Africa | $c_{e1u}$ | 0.29 | | 0.29 | | 0.29 | | 0.29 | | 0.29 | | 0.10 | 30 |
| South Africa | $d_1$ | 0.01 | | 0.0001 | | 0.0001 | | 0.0001 | | 0.0001 | | 0.25 | Assumption |
| South Africa | $d_2$ | 0.021 | | 0.00046 | | 0.00046 | | 0.00046 | | 0.00046 | | 0.25 | Assumption |
| South Africa | $d_2$ | 0.04 | 2020-07-01 | | | | | | | | | 0.25 | Assumption |
| South Africa | $d_2$ | 0.01 | 2020-07-25 | | | | | | | | | 0.25 | Assumption |
| South Africa | $d_m$ | 0.25 | | 0.0011 | | 0.005 | | 0.005 | | 0.005 | | 0.25 | Assumption |
| South Africa | $d_m$ | 0.32 | 2020-05-01 | | | 0.0031 | 2020-05-05 | 0.0031 | 2020-05-05 | 0.0031 | 2020-05-05 | 0.25 | Assumption |
| South Africa | $d_m$ | 0.5 | 2020-07-01 | | | 0.0008 | 2020-08-01 | 0.0008 | 2020-07-15 | 0.0008 | 2020-07-15 | 0.25 | Assumption |
| South Africa | $d_m$ | 0.1 | 2020-07-25 | | | | | 0.002 | 2020-08-01 | 0.002 | 2020-08-01 | 0.25 | Assumption |
| South Africa | $d_m$ | 0.05 | 2020-08-01 | | | | | 0.0032 | 2020-09-15 | 0.0032 | 2020-09-15 | 0.25 | Assumption |
| South Africa | $d_s$ | 0.25 | | 0 | | 0 | | 0 | | 0 | | 0.25 | Assumption |
| South Africa | $ddf$ | | | 0.2 | | 0.2 | | 0.2 | | 0.2 | | 0.25 | Assumption |
| South Africa | $ddf$ | | | | | 0.12 | 2020-08-01 | 0.12 | 2020-08-01 | 0.12 | 2020-08-01 | 0.25 | Assumption |
| South Africa | $\delta_{hadjust}$ | 1 | | 1 | | 1 | | 1 | | 1 | | 0.00 | Assumption |
| South Africa | $\delta_{hadjust}$ | 0.44 | 2020-04-30 | | | 0.9 | 2020-06-15 | 0.9 | 2020-06-15 | 0.9 | 2020-06-15 | 0.00 | Assumption |
| South Africa | $\delta_{su}$ | 0.0075 | | 0.0022 | | 0.0022 | | 0.0022 | | 0.0022 | | 0.25 | WHO mission |
| South Africa | $\delta_c$ | 0.04 | | 0.023 | | 0.023 | | 0.023 | | 0.023 | | 0.25 | 34 |
| South Africa | $\delta_h$ | 0.02 | | 0.0025 | | 0.0025 | | 0.0025 | | 0.0025 | | 0.25 | 34 |
| South Africa | $\eta_u$ | 0.18 | | 0.2 | | 0.2 | | 0.2 | | 0.2 | | 0.25 | 35–40 |
| South Africa | $h_{ceil}$ | 1E+10 | | 1E+10 | | 1E+10 | | 1E+10 | | 1E+10 | | 0.00 | Assumption |
| South Africa | $hdf$ | | | 0.28 | | 0.32 | | 0.32 | | 0.32 | | 0.25 | Assumption |
| South Africa | $hdf$ | | | | | 0.17 | 2020-08-01 | 0.17 | 2020-08-01 | 0.17 | 2020-08-01 | 0.25 | Assumption |
| South Africa | $r_{2u}$ | 0.097 | | 0.097 | | 0.097 | | 0.097 | | 0.097 | | 0.25 | 41 |
| South Africa | $r_{mu}$ | 0.096 | | 0.096 | | 0.096 | | 0.096 | | 0.096 | | 0.10 | 41 |
| South Africa | $r_h$ | 0.35 | | 0.35 | | 0.28 | | 0.28 | | 0.28 | | 0.25 | 34 |
| South Africa | $r_h$ | | | | | | | 0.33 | 2020-09-01 | 0.33 | 2020-09-01 | 0.26 | 34 |
| South Africa | $r_p$ | 0.3 | | 0.3 | | 0.3 | | 0.3 | | 0.3 | | 0.25 | Assumption |
| South Africa | $\rho$ | 0.12 | | 0.19 | | 0.19 | | 0.19 | | 0.19 | | 0.25 | 34 |
| South Africa | $\rho$ | | | | | | | 0.25 | 2020-09-01 | 0.25 | 2020-09-01 | | |
| South Africa | $\theta$ | 0.2 | | 0.022 | | 0.022 | | 0.022 | | 0.022 | | 0.25 | 35, 37, 42 |
| South Africa | FOIadjust | 1 |  | 1 |  | 1 |  | 1 |  | 1 |  | 0.00 | Assumption |
| South Africa | FOIadjust | 0.32 | 2020-03-31 | 0.35 | 2020-04-20 | 0.35 | 2020-04-20 | 0.35 | 2020-04-20 | 0.35 | 2020-04-20 | 0.25 | Assumption |
| South Africa | FOIadjust | 0.4 | 2020-05-05 | 0.4 | 2020-05-15 | 0.36 | 2020-05-15 | 0.36 | 2020-05-15 | 0.36 | 2020-05-15 | 0.25 | Assumption |
| South Africa | FOIadjust | 0.28 | 2020-06-14 | 0.35 | 2020-06-21 | 0.34 | 2020-06-15 | 0.35 | 2020-06-15 | 0.35 | 2020-06-15 | 0.25 | Assumption |
| South Africa | FOIadjust | 0.11 | 2020-07-30 | 0.12 | 2020-07-16 | 0.18 | 2020-07-15 | 0.13 | 2020-07-20 | 0.13 | 2020-07-20 | 0.25 | Assumption |
| South Africa | FOIadjust |  |  | 0.25 | 2020-09-01 | 0.25 | 2020-08-25 | 0.25 | 2020-09-05 | 0.25 | 2020-09-05 | 0.25 | Assumption |
| South Africa | FOIadjust |  |  |  |  |  |  | 0.29 | 2020-10-01 | 0.29 | 2020-10-01 | 0.25 | Assumption |
| South Africa | FOIadjust |  |  |  |  |  |  | 0.45 | 2020-11-01 | 0.57 | 2020-11-10 | 0.25 | Assumption |
| South Africa | FOIadjust |  |  |  |  |  |  |  |  | 0.56 | 2020-12-15 | 0.25 | Assumption |

**Table 4.** Description of model parameters

| Parameter | Description |
| --- | --- |
| $a_{1d}$ | Relative infectivity of diagnosed, pre-symptomatic people $I_{1d}$ |
| $a_{1u}$ | Relative infectivity of undiagnosed, pre-symptomatic people $I_{1u}$ |
| $a_{2d}$ | Relative infectivity of diagnosed, asymptomatic people $I_{2d}$ |
| $a_{2u}$ | Relative infectivity of undiagnosed, pre-symptomatic people $I_{2u}$ |
| $a_{md}$ | Relative infectivity of diagnosed people with mild symptoms $I_{md}$ |
| $a_{sd}$ | Relative infectivity of diagnosed people with severe symptoms $I_{sd}$ |
| $a_{su}$ | Relative infectivity of undiagnosed people with severe symptoms $I_{su}$ |
| $b_b$ | Effective contact rate, which encompasses all of the biological and behavioral considerations that influence contacts between individuals that lead to transmission |
| $c_{12u}$ | Inverse of the average stay in the $I_{1u}$ state for undiagnosed and asymptomatic people |
| $c_{1mu}$ | Inverse of the average stay in the $I_{1u}$ state for undiagnosed and mildly symptomatic people |
| $c_{1su}$ | Inverse of the average stay in the $I_{1u}$ state for undiagnosed and severely symptomatic people |
| $c_{ceil}$ | This is the maximum number of people who can be in critical care facility at a single time step. |
| $c_{e1u}$ | Inverse of the average stay in the $E_u$ state (undiagnosed and infected but not yet infectious) |
| $d_1$ | Inverse of the average time for people in the $I_{1u}$ state (pre-symptomatic) until they get diagnosed with coronavirus infection, while still pre-symptomatic |
| $d_2$ | Inverse of the average time for people in the $I_{2u}$ state (infectious but asymptomatic) until they get diagnosed with coronavirus infection, while still infectious and asymptomatic |
| $d_m$ | Inverse of the average time for people in the $I_{mu}$ state (mildly symptomatic) until they get diagnosed with coronavirus infection, while still infectious and mildly symptomatic |
| $d_s$ | Inverse of the average time for people in the $I_{su}$ state (severely symptomatic) until they get diagnosed with coronavirus infection, while still infectious and severely symptomatic |
| $ddf$ | This is the fraction of deaths that happened outside of the hospital where COVID-19 is identified as the cause of death |
| $\delta_{su}$ | Inverse of average time till death for people with severe symptoms $I_{su}$ who die without any hospitalisation |
| $\delta_c$ | Inverse of average time till death for people who die while admitted to critical care |
| $\delta_h$ | Inverse of average time till death for hospitalised people who die without any stay in critical care |
| $\delta_{hadjust}$ | This is the factor by which you multiply the $\delta_h$ parameter if you want to decrease the rate at a specific point in time |
| $\eta_u$ | Inverse of average time till hospitalisation for undiagnosed people with severe symptoms $I_{su}$ |
| $h_{ceil}$ | The maximum number of people who can be in the hospital at a given time |
| $hdf$ | The fraction of all people who go to the hospital with COVID-19 that are actually diagnosed with COVID-19 |
| $r_{2u}$ | Inverse of the average stay in the $I_{2u}$ state (asymptomatic, undiagnosed) |
| $r_{mu}$ | This is the rate at which undiagnosed mildly infected individuals recover |
| $r_h$ | This is the rate at which hospitalised individuals recover, without going to critical care |
| $r_p$ | This is the rate at which individuals in the post-critical care ward recover |
| $\rho$ | This is the rate that those in the critical care ward move to the post-critical care ward |
| $\theta$ | This is the rate that those who are hospitalised move to the critical care ward |
| FOIadjust | This is the factor by which you multiply the force of infection if you want to decrease the rate at a specific point in time |

### Model equations

The model is formally represented by a system of differential equations:

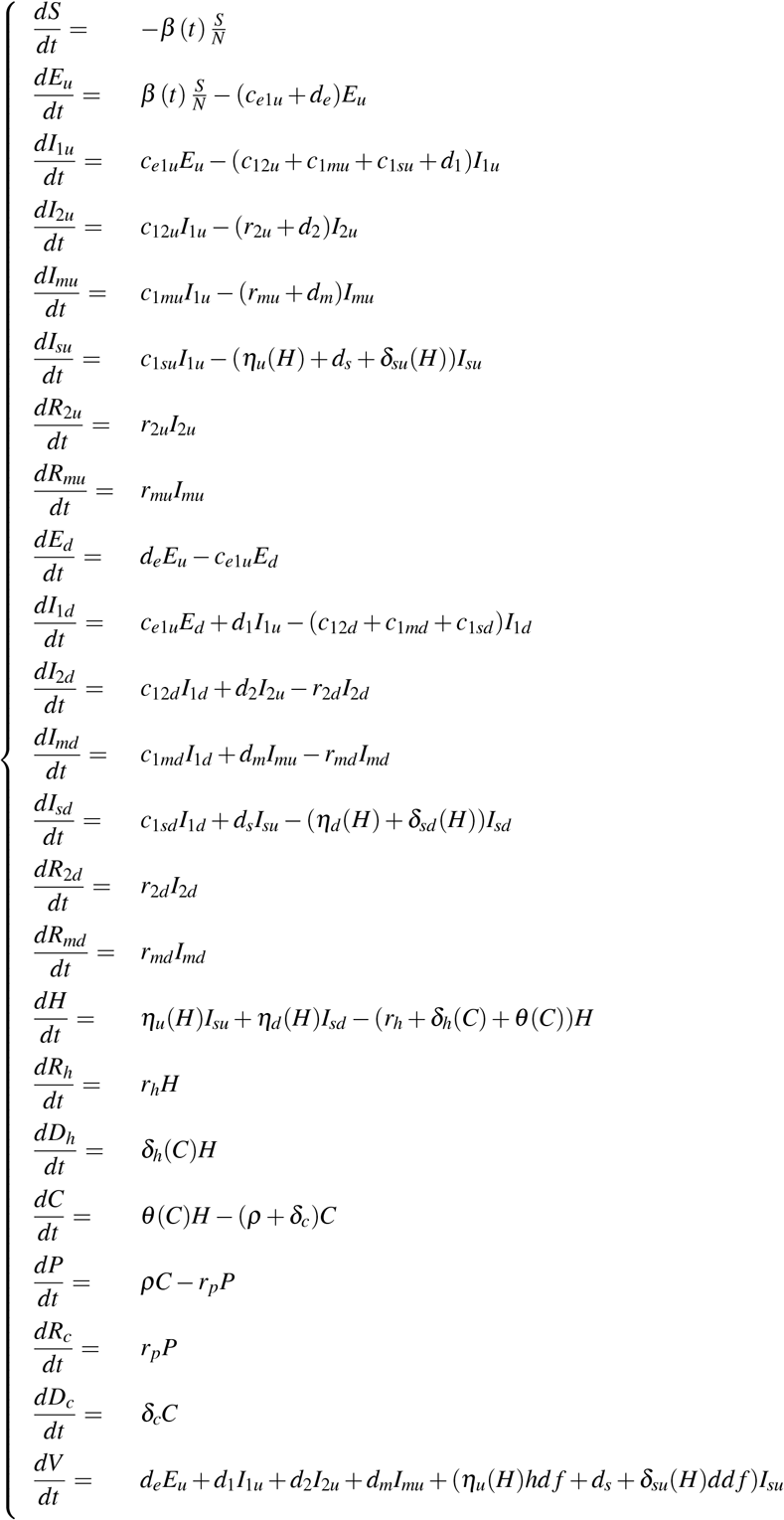

with

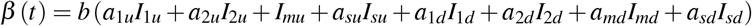

and *V* representing the number of confirmed cases.

### Hospital and critical care capacity limitations

The capacity for hospitals and critical care units to admit patients is not unlimited in South Africa - as in most countries. Defining *v* as the hospital capacity limit, the effective rate of hospitalisation is: *ηH* while *H* < *v*. When *H* starts to exceed *v*, it becomes 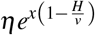, which is ≈ 0 for large values of *x*. Similarly, the effective rate of critical care admission is: *θC* while *C* < *w*. When *C* starts to exceed *w*, it becomes 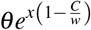.

### Basic reproductive number

The basic reproductive number *R*_0_ is given by

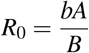

where

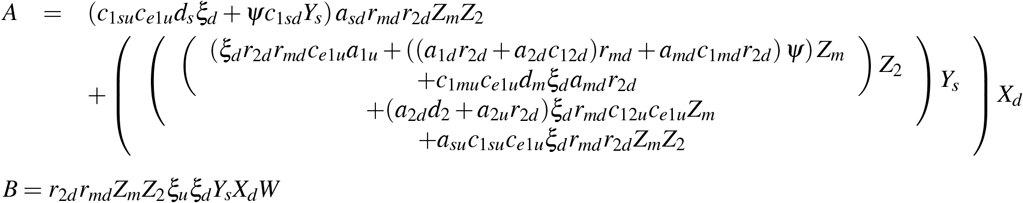

with

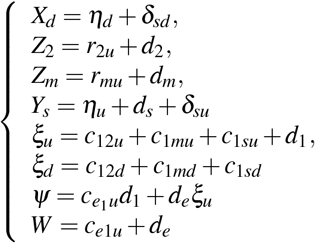

### Effective reproductive number

The effective reproductive number *R*_*e*_ (*t*) is given by

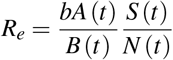

where

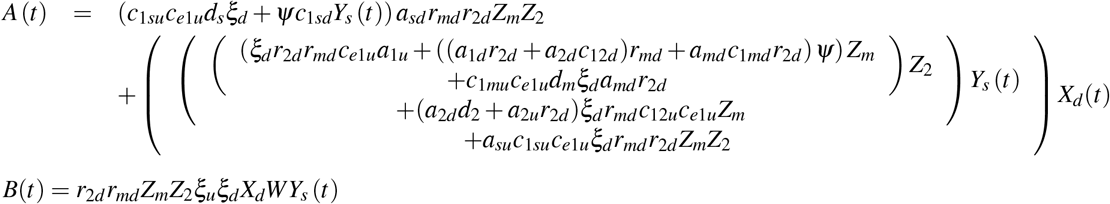

with

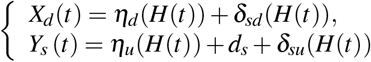

